# Structural Variants Are a Major Component of the Missing Heritability of Autism Spectrum Disorder

**DOI:** 10.1101/2021.10.10.21264819

**Authors:** David Kainer, Alan Templeton, Erica T. Prates, Euan Allan, Sharlee Climer, Daniel A. Jacobson, Michael R. Garvin

**Author notes:** These authors contributed equally (David Kainer, Michael R. Garvin). This manuscript has been authored by UT-Battelle, LLC under Contract No. DE-AC05-00OR22725 with the U.S. Department of Energy. The United States Government retains and the publisher, by accepting the article for publication, acknowledges that the United States Government retains a non-exclusive, paid-up, irrevocable, world-wide license to publish or reproduce the published form of this manuscript, or allow others to do so, for United States Government purposes. The Department of Energy will provide public access to these results of federally sponsored research in accordance with the DOE Public Access Plan (http://energy.gov/downloads/doe-public-access-plan).

## Abstract

The heritability of autism spectrum disorder (ASD), based on 680,000 families and five countries, is estimated to be nearly 80%, yet we lack genetic markers that adequately explain it. It is increasingly clear that genomic structural variants (SVs) are a major component of the “missing heritability” for many complex phenotypes. Here we use a novel method to identify SVs based on non-Mendelian inheritance patterns in pedigrees using parent-child genotypes from ASD families and demonstrate that the genes that the ASD-specific SVs overlap recapitulate the known molecular biology of ASD including dendritic spinogenesis, axon guidance, and chromatin modification. We further define fine-grained biological pathways that strongly implicate aberrant early development of the cerebellum. Importantly, using these previously excluded variants, we identify the *ACMSD* gene in the kynurenine pathway as significantly associated with non-verbal cases of ASD and we then use an explainable artificial intelligence approach to define subgroups for future diagnosis and deployment of personalized medicine.

## Introduction

Although the heritability of autism spectrum disorder (ASD) is nearly 80% ^1^, only a small proportion can be explained by single nucleotide polymorphisms (SNP) driven Genome Wide Association Studies (GWAS) ^2^. This may be partly explained by a lack of statistical power as ASD is phenotypically and molecularly heterogeneous (i.e., mutations in different components of a biological process or processes underlie the phenotype), and thus requires large sample sizes to detect variants of smaller effect. Alternatively, it is possible that the “missing heritability” is in the form of genomic structural variants (SVs, genomic alterations larger than 50 base pairs) of large effect that have not been included in previous analyses.

It is becoming increasingly clear that SVs are a major component of missing heritability in complex traits including human disease ^3^, which is not surprising given that they affect approximately five times the amount of genomic space, are three times more likely to be associated with a GWAS signal, and are fifty times more likely to affect the regulation of a gene ^4^ compared to SNPs. For example, amyotrophic lateral sclerosis (ALS), like ASD, is a heterogeneous disorder with an estimated heritability of 65%, and yet large-scale genomic analyses have only identified markers that explain about 10% of cases. However, it is now known that SVs caused by expansion of repetitive microsatellite elements in two genes (*C9orf27* and *ATXN2*) cause some cases of ALS ^5^. Likewise, the heritability of late onset Alzherimer’s disease (LOAD) is at least 60%, and although the epsilon 4 allele of *ApoE* accounts for roughly a quarter of that heritability, it does not fully explain age of onset or the remaining cases ^6^. However, an SV in the neighboring gene *TOMM40*, which likely represents a hotspot for transposon activity ^7^, increases the LOAD risk odds ratio by 4-fold compared to the *ApoE e4* allele alone ^8^. SVs are a known risk factor for developmental disorders such as ASD ^2,9^, but as with SNPs, there are currently none that are consistently replicated in large numbers of ASD cases.

A major reason that SVs are typically excluded from GWAS ^10^ is that their detection is prone to false positives and low recall ^4^, particularly from short-read sequencing data ^11^. However, a 2006 study by Conrad et al demonstrated an elegant method for detecting large SVs (deletions) from SNP array genotyping of parent-child trio ^12^. During the array-based genotyping process, an SV may cause SNPs at or near the locus to show patterns of non-mendelian inheritance (NMI) and non-conformance to Hardy-Weinberg expectations (HWE). The underlying SV itself is likely segregating like any polymorphic site in the genome, but when parent and child have a genomic disruption at a locus, it causes the hybridizing probe to bind the target incorrectly (**Fig. 1a**); in these situations, genotyping algorithms interpret the signal as invalid and do not provide a call. If the SV is heterozygous this often results in an NMI pattern. Conrad et. al. showed that SNPs exhibiting NMI patterns can therefore be used as a proxy for SV discovery. Interestingly, such NMI SNPs are almost universally filtered out at the quality control stage of GWAS under the assumption that they represent erroneous genotyping, thereby potentially removing a critical component of the heritability of the trait.

**Fig. 1.**
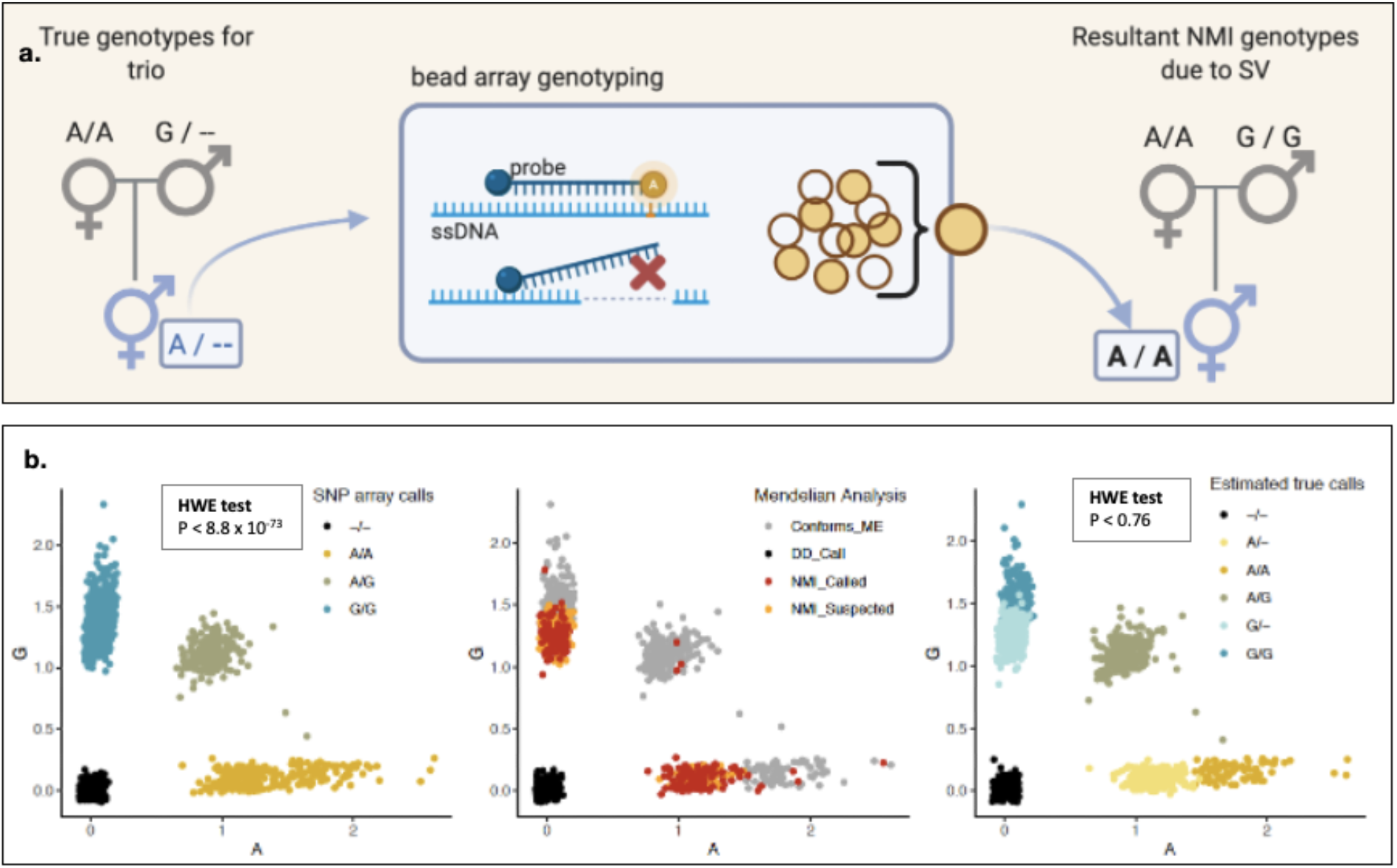
Non-Mendelian Inheritance (NMI) to detect normally segregating SVs. (**a**) an NMI signal can occur when an SV exists under the region of DNA that is targeted by the hybridizing probe (red “X”). In this example scenario, the missing signal from one allele coupled with a normal signal from the other allele produces an erroneous genotype (pedigree on the right) that does not conform to mendelian expectation of the trio. (**b**) For example, array genotyping of the ASD trio children for SNP *rs221465* results in failure of the Hardy-Weinberg expectation (HWE) test (left). PLINK *mendel* reveals many individuals with NMI (center plot, red dots) at this SNP. However, there are further individuals where we “suspected NMI” (center, orange dots). These individuals are from trios where PLINK had no power to detect NMI as all three individuals were genotyped as A/A, but they co-locate with the NMI individuals on the signal intensity plot. We inferred the genotype calls for NMI and suspected NMI cases (right), and now this SNP conforms to HWE (note that point locations between plots vary slightly due to an applied jitter). Indeed it is already known that this SNP tags a large common deletion in the *NRXN3* gene. The allele frequency of the deletion (“-”) in the ASD population after the NMI-based correction (0.34) is highly similar to the frequency in the 1000 Genome population (0.37).

Here we extend the use of NMI for SV detection beyond the original approach of Conrad et al. Whereas the initial method detected only deletions, we recognized other complex and important SVs, including copy number gains, can also produce an NMI pattern. For example, deletions will decrease signal intensities for an allele whereas duplications will cause an increase. The important feature is that a SNP may display NMI if it does not fall within the three expected intensity clusters defined by homozygous for allele 1, homozygotes for allele 2, or heterozygotes (**Fig. 1b**). When a SNP falls outside those boundaries, the software issues a “no call” or an incorrect call even though there is an intensity signal. Furthermore, the power of a pedigree allows one to determine if the SV is maternally or paternally inherited, and unlike Conrad et al., we can score a potential heterozygous SV in a child based simply on those that are marked as “no calls” in the parent (i.e. homozygous SVs in a parent that can be scored in the offspring). Finally, the availability of a highly annotated human genome (reference human genomes hg17 for Conrad et al. compared to the current hg38) and much higher SNP densities provides an opportunity to integrate SV calling with a systems biology approach to determine if these represent missing heritability in complex traits.

Here, rather than discarding such SNPs, we explicitly identified SNP loci that displayed patterns of NMI (i.e. NMI-SV) in one ASD parent-child trio dataset ^13^, validated them in a second dataset ^14^, and then applied stringent filters to remove any potential false positives. The remaining loci recapitulate the ASD phenotype and confirm previous molecular studies. For example, infrequent but statistically significant variants in *SYNGAP1* have been reported as associated with ASD in more than 50 studies (sfari.org); here, we identify an NMI-SV in 17% of ASD cases that overlaps a reportedly statistically significant GWAS SNP site (rs9461856). Likewise, in nearly one-third of ASD cases we detect a highly significant paternally transmitted NMI-SV at the SNP rs11739167 located in the *MSNP1* gene. This is within 60 kb of SNP rs4307059, which was reported as associated with ASD in one of the source studies we used ^15^, but did not pass significance threshold testing in that work. Aside from confirming previously reported sites but with greater significance, we tested to determine if genes marked by NMI-SV significantly over-represent unique biological processes and found that they are in fact enriched for the known ASD-associated processes of dendritic spinogenesis, glutamate receptor signaling, chromatin modification, as well as axon guidance regulated by neural cell adhesion molecules and guidance cues. By pinpointing high-frequency SVs (within the 50bp span of the genotyping probe), we were able to reveal how these processes are linked and that they appear to be important for early development of the cerebellum and telencephalon.

Finally, and most importantly, we performed an association test using only ASD-specific NMI-SV (ASD-SV) and identified a significant association with non-verbal cases in a transcription factor binding site that regulates the *ACMSD* gene in the tryptophan catabolism pathway, confirming that these variants represent at least some of the missing heritability in ASD. We then apply an explainable artificial intelligence (XAI) algorithm to the matrix of SVs to identify subgroups of ASD individuals and the affected genes that define them.

## Results

### SV detection and filtering

We performed NMI tests in PLINK on both the MIAMI and AGPC datasets, which flagged 101,032 putative SV sites (i.e., having at least one family with NMI in one or both of the data sets). We then manually scored these 101,032 sites for NMI in further families that PLINK did not flag and estimated the frequencies within each population (**Fig. S1, Fig 2**). Out of a total of 338.4m genotyped sites in the MIAMI data set (i.e., 380 children × 890,539 SNPs used), 1.23m displayed an NMI pattern, or 0.36% of total genotyping assays across the 380 arrays.

**Fig. 2.**
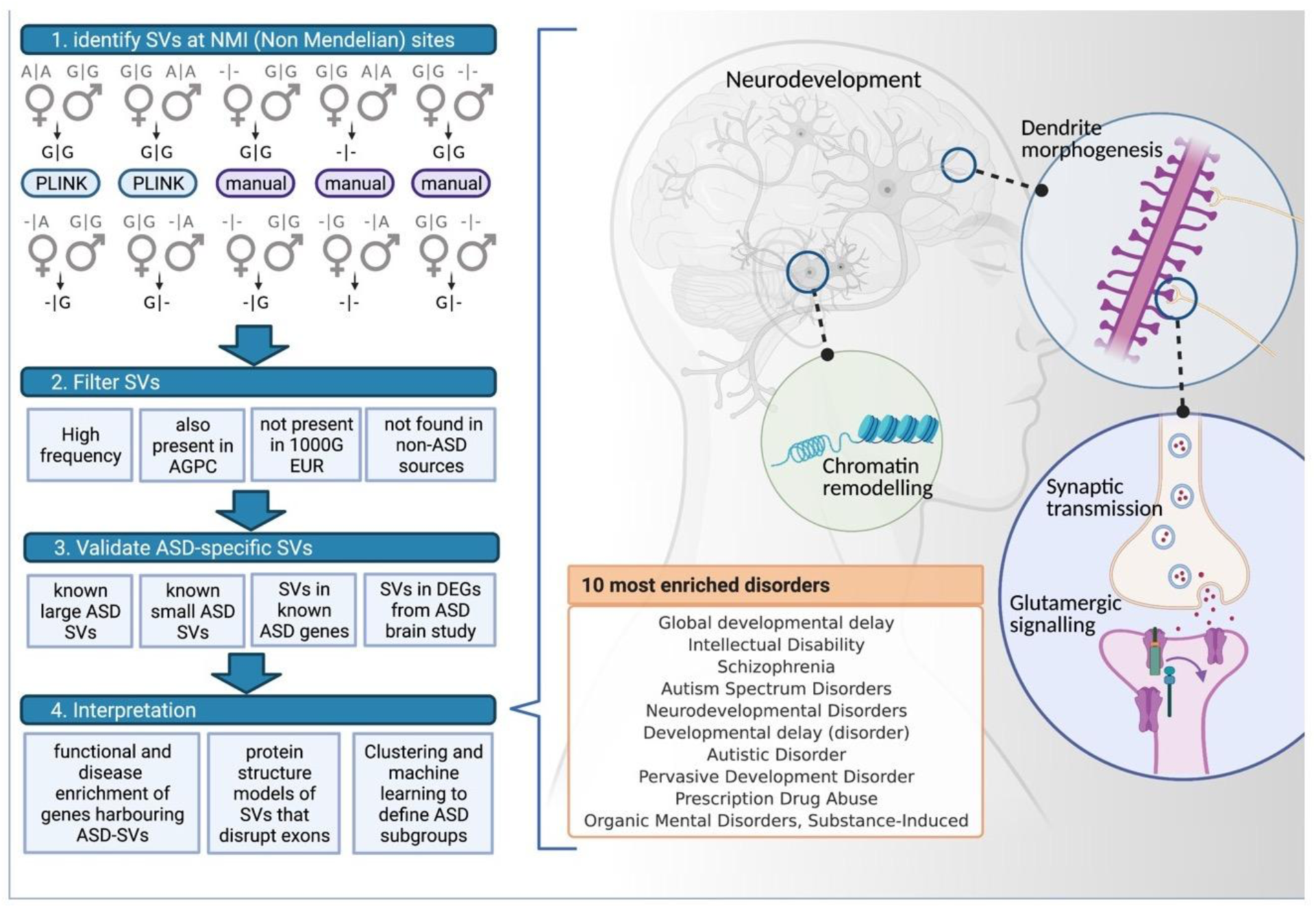
Detection, filtering, and functional enrichment of ASD-SV. NMI is used to identify potential SVs from parent-child trios (left), either with PLINK or manually. A set of filters are then applied, including removing SVs found in non-ASD studies, and the remaining SVs are subjected to several validation processes and downstream analyses. Coding genes that harbored ASD-SVs marked by NMI SNPs found at greater than 15% frequency in both study populations were assessed for enrichment of GO Biological Process terms (right). Four major categories were found to be statistically significant (false discovery rate, FDR < 0.05, fold-enrich > 2, **Table S6a**):, dendritic morphogenesis, neuronal migration, synaptic transmission, and glutamate receptor signaling. The genes also display significant enrichment in ASD and other neurodevelopmental disorders using the disease ontology association in ToppGene Suite (ToppGene.cchmc.org) as well as in transcription factor binding sites involved in chromatin remodeling. A GO analysis of the 118 genes that contain ASD-SV_NMI_ and overlapping with recently reported regulatory regions for development of the telencephalon (**Table S7**) returned nearly identical significantly enriched processes.

After removing rare SVs with a frequency less than 2% in the MIAMI population, we were left with 61,703 as our discovery panel. Of these, 55,767 (90%) were also detected as SVs in at least one family in the AGPC population (**Fig. 3a**, our QC determined that no individuals were present in both data sets, ***Supp Methods***). This set was labeled as NMI-SV. The frequencies of the discovery SVs in MIAMI were strongly correlated with those in AGPC (Pearson’s r=0.75; p < 0.0001), supporting the accuracy of this approach. To obtain the ASD-specific set of SVs we next removed NMI-SV that were previously reported and known SVs from several sources including the 1000 Genome Project (1KGP, **Fig S1, Fig 2**, see Methods). This left a total of 48,009 SVs in the ASD-SV set (5.5% of all sites in the array that passed QC) with frequency greater than 2% in the MIAMI population (**Table S1, S2**). The core of the ASD-SV set was defined by 1,175 SVs with greater than 15% frequency in both the MIAMI and AGPC populations, located in 1,106 protein-coding genes (**Fig. 3b**). On average, each individual in AGPC had 371 coding genes harboring high frequency ASD-SVs, while individuals in MIAMI averaged 347 (**Fig 3c**).

**Fig. 3.**
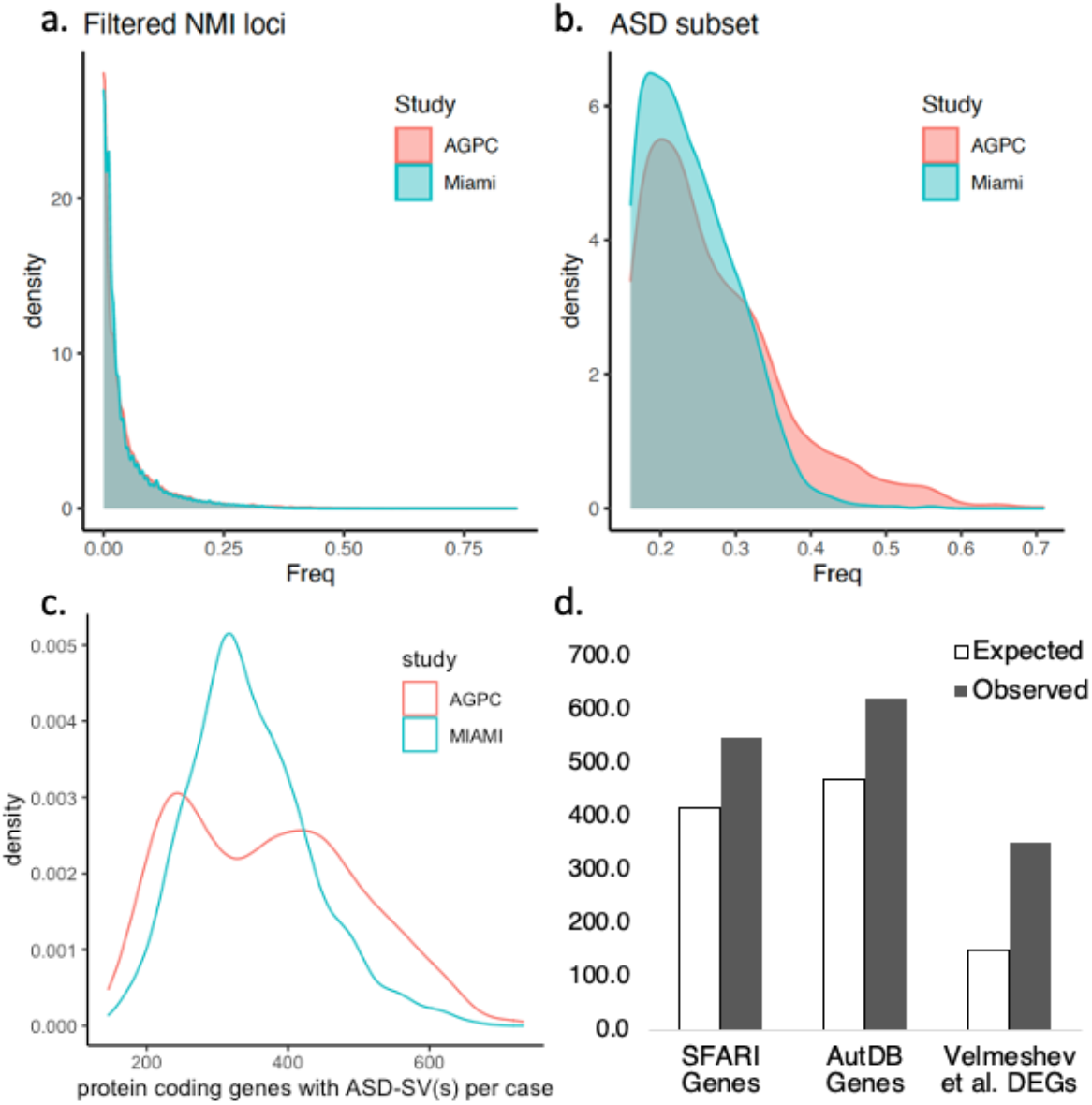
(**a**) NMI patterns identified over 60,000 likely structural variants (NMI-SV) in the smaller MIAMI data set (blue) and the vast majority (90%) were validated in the larger AGPC data set (pink) with a very similar frequency spectrum. Removal of known SVs from non-ASD populations left 48,009 ASD-specific SVs (ASD-SVs), most of which were rare. (**b**) There is a considerable overlap of the highest frequency ASD-SVs between the two studies (right) indicating a likely core set of SVs underlying ASD. (**c**) Density distributions of the number of genes with high-frequency ASD-SVs per individual. This was done separately for the AGPC and Miami cohorts. The number of genes harboring ASD-SVs varies per case, potentially determining the spectrum of ASD phenotype. On average, each individual in AGPC had 371 genes harboring high frequency ASD-SVs, while individuals in MIAMI averaged 347 (**d**) NMI-SVs identify more known ASD genes than is expected by chance in the SFARI (p < 1 × 10 ^-10^) and AutDB (p < 3.6 × 10 ^-12^) data sets and in the recently reported differentially expressed genes in post-mortem brain tissue of ASD individuals (p < 3 × 10 ^-60^) ^17^.

### The NMI method strongly recalls known ASD-related SVs

The SVs most confidently identified using the NMI method are those that represent large deletions that span multiple contiguous (on a chromosome) SNPs. The SNP loci are randomized on the array and therefore the probability of seeing NMI at each of these genomically contiguous SNPs by chance is extremely low. For example, we identified NMI at 43 contiguous, physically linked SNPs in three individuals in the MIAMI data set (**Fig. S2**). Based on the overall NMI rate across the array, the probability of finding this number of physically adjacent NMI loci due to technical error is exceedingly small (1.2 × 10^−105^) (**Supplementary Methods**). Indeed, this particular stretch of 43 NMI SNPs most likely identifies a large SV that is known to cause subtypes of ASD including Angelman Syndrome ^16^ (**Fig. S2b**). By using these high-confidence consecutive NMI-SVs we were able to identify 15 of the 17 ASD-susceptibility loci that are known to be large chromosomal disruptions (**Table S3**).

To further test our approach, we examined the SNPs that overlapped known ASD-associated copy number variation (CNV) SVs. The Autism DataBase (AutDB) lists CNV identified from the 28,735 ASD cases. Of the 2,270 small CNVs from AutDB that were potentially detectable with the SNPs on the Illumina array, our NMI approach captured 1,902 (84%) of them (**Table S4**). This is a challenging test, since small CNVs overlap only one or two SNPs. Therefore, the result is highly supportive of the efficacy of NMI as a proxy for CNV detection.

### ASD susceptibility genes are more likely to harbor NMI-SVs

Of the 16,917 protein coding genes marked by the sites on the Illumina array, 49% (8,222) had at least one NMI-SV associated with them. The SFARI database lists 1,003 ASD risk genes (see Data Description and Methods), of which 866 are marked by the Illumina array used in the MIAMI and AGPC studies. Assuming a random distribution of NMI-SVs across the genome, our expectation was that 421 of these genes would harbor an NMI-SV. However, we found NMI-SVs in a significantly greater number (600, or 69%); (*chi*-square test p < 2.5 × 10^−18^; **Fig. 3d, Table S5**). Likewise, AutDB lists 1,241 ASD risk genes, of which 1,072 are marked by the array used here. We would expect to find 521 genes harboring NMI-SVs but, instead, we find a significantly greater number (n=748, p < 2.7 × 10^−23^, *chi*-square test, F**ig. 3d, Table S5**).

### NMI identifies SV that explains the original MSNP1 GWAS signal

The parent-child trio studies from which we obtained our data were initially designed to detect transmission disequilibrium. For example, an inherited maternal deletion at 15q11.2 can cause Angelman Syndrome (AS) but a paternally inherited deletion at this region causes Prader-Willi-Syndrome (PWS); the two disorders are phenotypically very different, which is likely a result of the paternal and maternal alleles being expressed at different developmental timepoints. The studies that generated the data we analyzed here ^14,15^ hypothesized that ASD results from differences in maternal or paternal inheritance at SNPs. Although they did not report any significant differences,, using a transmission disequilibrium test (TDT), we identified ten ASD-SV that displayed significantly greater than expected maternal inheritance and four that showed greater than expected paternal inheritance (**Fig. 4a** and **Table S6**), in line with the SV patterns observed for AS and PWS.

**Fig. 4.**
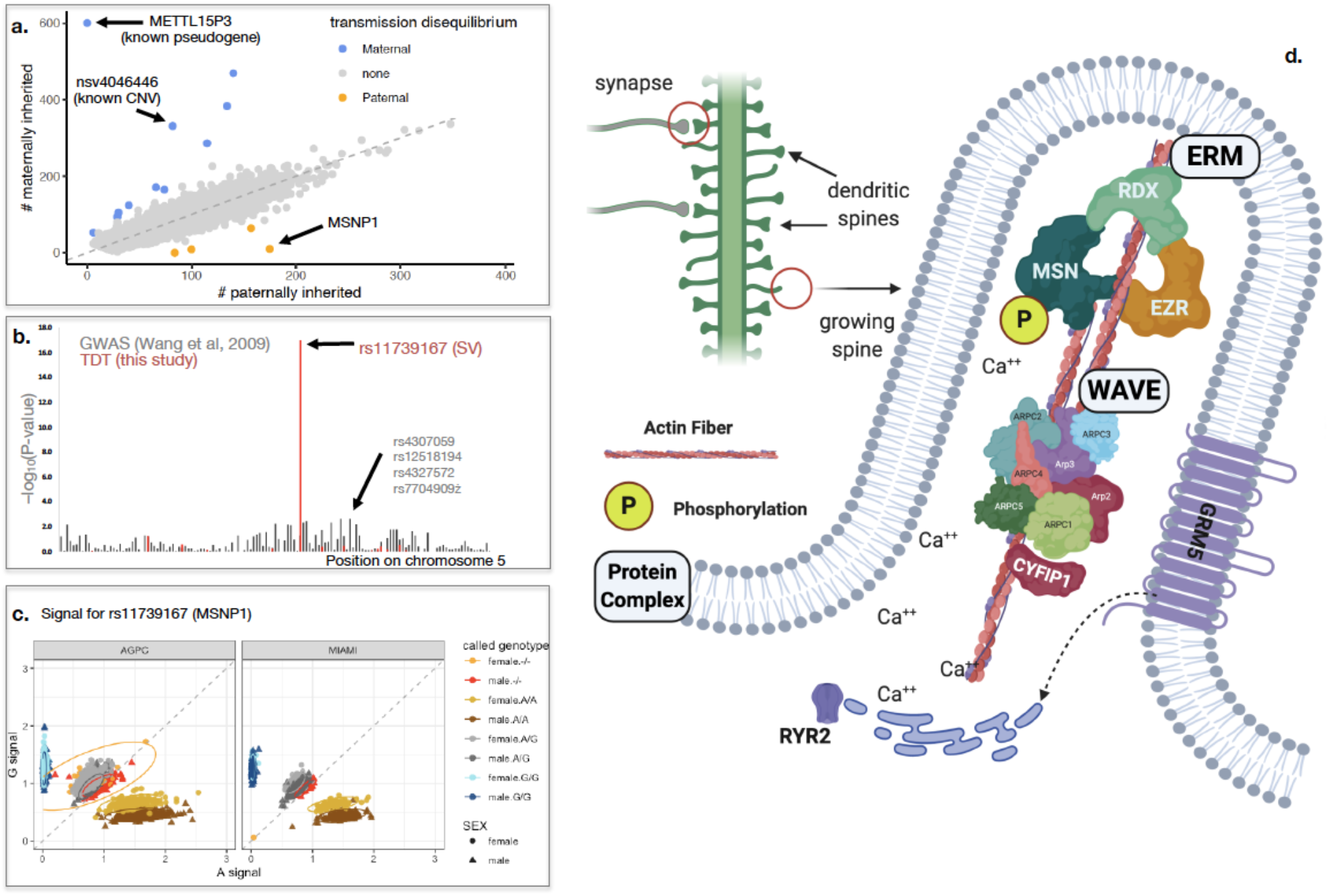
Significant transmission disequilibrium of ASD-SV. (**a**) A plot of ASD-SV reveals several loci that are maternally (blue) and paternally (orange) over-transmitted (i.e., significantly different than freq = 0.50). The maternal SV (*y*-axis) is truncated to 450 individuals for clarity. There was a nearly 18-fold greater than expected number of SV_NMI_ tagged by the rs11739167 SNP in the *MSNP1*. **(b**) The *MSNP1* ASD-SV is roughly 60 kb from a SNP strongly associated with ASD in Ma et al. 2009. This SNP met significance after imputing and combining several data sets in a second study (Wang et al. 2009). Here, the *p*-values for those SNPs were determined based on the allele frequencies of the ASD genotypes in the cases and the 1KGP controls. *MSNP1* is a pseudogene that regulates the expression of Moesin (*MSN*). (**c**) A plot of the original intensity values for the A (*x*-axis) and G (*y*-axis) alleles at the *MSNP1* SNP rs11739167 reveals that the original call of “no signal, -/-” is actually a subgroup of different alleles in fathers and the male offspring. The vertical shift of intensity values for all ASD females and mothers (orange triangles) is most likely probe hybridization to the parent gene on the X chromosome. In contrast, an increase in intensity of a subset of males on the *x*-axis indicates a CNV in that subgroup on the autosome (chromosome 5). The same pattern is present in the Miami data set. (**d**) MSN is activated by phosphorylation *via* GRM5 (also called mGluR5) glutamate receptor signaling and ryanodine-mediated calcium release from the endoplasmic reticulum. MSN is a component of a complex that manipulates the actin cytoskeleton to develop dendritic spines, which are the major form of learning and plasticity in the developing brain and are altered in ASD ^30^. Another major component of this process is the WAVE complex, which includes the CYFIP1 protein that was identified in our large deletion (**Fig. S2**) as well as the Fragile-X-causing FMR1 gene. EZR=Ezrin, RDX=Radixin, RYR2=Ryanodine Receptor 2.

The most significant paternally transmitted ASD-SV (spanning rs11739167, 17.6-fold greater than expected based on a *χ* -square test, *p* < 9.7 × 10^−18^; **Fig. 4a**) is centered on the Moesin Pseudogene 1 (*MSNP1*), a non-coding RNA that regulates its parent gene (*MSN*) on the X chromosome ^18^. Crucially, this ASD-SV is centered on the primary GWAS signal of the original study from the Miami data set and a subsequent study that incorporated those individuals ^13,15^ (**Fig. 4b**) but was removed from that analysis because it did not conform to Mendelian inheritance (**Supplementary File 1**). Expression of *MSNP1* was shown to be significantly greater in post-mortem brain tissue of ASD individuals who are homozygous for the T allele at the nearby SNP rs4307059 compared to controls (**Fig. 4b**) ^19^. Additionally, five large CNVs in this region were validated with PCR ^13^ and we were able to detect all of them, indicating that our method is easily able to identify the strongest signals from Wang *et al*. with substantially increased significance (**Table S6b**).

A re-plot of the genotypes using the raw signal intensities for the parent-child trios at the rs11739167 SNP in *MSNP1* (**Fig. 4c**) at first glance appeared to indicate that the NMI signal was a result of cross-hybridization of the *MSNP1* probe with the *MSN* parent gene on the X chromosome. However, upon closer inspection, one can see that the subset of males displaying the ASD-SV pattern are shifted to the right and *not* vertically. This indicates a change on chromosome 5. In contrast, in females, the probe binding to the *MSN* parent gene on the X chromosome produces an upward shift of all individuals on the *y*-axis and this does *not* occur in males. This is because the site on the X chromosome is invariant, i.e., only produces a ‘G’ allele, whereas the site in the *MSNP1* gene on chromosome 5 is variable, producing an A or G. The increased intensity is consistent with a copy number gain with the A allele of *MSNP1* in a subset of males. Plots of the ASD offspring from the Miami study show the same pattern.

The remaining loci positive for the TDT also indicate that NMI-SV are able to distinguish between CNVs and pseudogenes associated with sex chromosomes. The NMI-SV at rs2024145 in the *METTLP2* pseudogene is likely due to probe binding to a copy (*METTLP3*) on the Y chromosome, which causes all males to shift upwards on the *y*-axis (**Fig. S3**). The NMI-SV TDT SNP rs3856834 identifies a known CNV (nsv4046446) on the Y chromosome and is found at roughly the same frequency in the 1KGP as it is in the ASD population here, indicating, as with *NRXN3* (**Fig. 1b**), it is likely not relevant to ASD, but is another example that our NMI method is accurately identifying SVs. A second NMI-SV SNP 2 kb away, rs10865745, appears to identify the same CNV (**Fig. S3**). Lastly, the NMI-SV at SNP rs10164308 appears to identify an active transposon (*HERVK11*; **Fig. S3**), which is now known to be a more common source of SVs than previously thought ^11^. The relevance of these NMI-SV to ASD are not yet known. In contrast, several independent lines of evidence implicate *MSNP1* in ASD, as we discuss next.

### MSNP1 implicates dendritic spines as central to ASD

Although early studies of the ASD-associated region on chromosome 5 originally focused on flanking cadherin gene ^13^, subsequent work has determined that the causative gene is most likely *MSNP1*. Expression of the pseudogene reduces MSN protein levels *in vitro*, which causes decreased neurite outgrowth in neural progenitor cells ^20^. Overexpression of *MSNP1* and closely linked enhancers in mice indicates it is tightly controlled and expressed during development of cerebral cortex and cerebellum ^21^. Disruption of *MSNP1* as a causative agent of ASD is also biologically cogent and consistent with previous reports. A GWAS identified *MSNP1* along with *PTPRD* and *GRIK2* as strongly associated with Obsessive Compulsive Disorder (OCD) ^22^, which is highly comorbid in mild forms of ASD ^23,24^, and which we implicate further in the following sections.

The parent gene *MSN*, which is regulated by *MSNP1*, encodes a component of the multi-protein complex ERM (ezrin/radixin/moesin) whose major function is to link cytoskeletal proteins to the membrane. In neurons, the ERM and WAVE protein complexes are central to the formation of dendritic spines ^25^ (**Fig. 4c**), which are short protrusions that extend from the main trunk of a dendrite. Their shape and size can change rapidly in short periods of time and they play a central role in early brain development, neural plasticity, and long-term memory ^26^. Dysfunction of dendritic spines have been thoroughly described in ASD ^27^.

Our results indicate that the atypical dendritic spine formation seen in some ASD individuals may be the result of a paternally inherited SV in the *MSNP1* pseudogene that causes dysregulation of the parent *MSN* gene. This is further supported by the fact that disruptions of other components of the WAVE complex also demonstrate aberrant dendrite spine morphology (e.g., FMRP is associated with Fragile X Syndrome and CYFIP1 with AS, as shown in our NMI analysis - **Fig. 4c, Fig. S2**) ^27^. MSN is activated when phosphorylated ^28^, which appears to mechanistically occur via intracellular calcium release mediated by ryanodine receptors from the endoplasmic reticulum that is in turn modulated by the glutamate receptor GRM5, or mGluR5 (see below) ^29^.

### Biological Functions of ASD-SV linked genes

#### Dendritic spines

Recent in-depth SV detection reports indicate there are roughly 28,000 SVs per individual in the human population ^4^. We found that each ASD case had, on average, several hundred coding regions of the genome containing one or more high frequency ASD-specific SV (Miami = 347, AGPC = 371; Fig. 3c). Given the stringent filtering of the initial NMI-SVs, their validation in a second independent ASD dataset, and their high recall of known ASD-related SVs, these SVs are likely a key component of the spectrum of ASD. A GO enrichment analysis of coding genes that harbor the core ASD-SVs revealed significant enrichment of biological process terms involved in dendritic spinogenesis, glutamate signaling, synaptic organization, and neuronal migration. All have been repeatedly linked to ASD ^31–34^, supporting the hypothesis that these NMI-derived SVs represent a major component of the missing heritability of the disorder and is consistent with the heterogeneity of ASD because they indicate disruption of multiple components of a few different biological processes. In addition, because our method identifies narrow regions of the genome that are affected, the resulting gene set is of high-confidence and uncovers previously unknown links between these processes as well as an expanded set of genes that underlie the disorder.

It is clear from these analyses that the set of core ASD-SVs, obtained via our NMI workflow in a cohort of ASD trios, contains a strong neurobiological signal, and not by random chance. While previous ASD reports have identified many of the biological processes we detected, only a handful of genes were attributed to these processes, and their seemingly diverse functions were attributed to pleiotropy ^35^. In contrast, here we find subgroups of genes that define fine-grained biological networks within these processes and, more importantly, functional linkages amongst them that indicate that these seemingly functionally diverse genes actually converge on the central process of **dendritic spine development in the cerebellum**. Our method also increases the number of genes associated with these biological pathways by nearly four-fold, further supporting the hypothesis that these loci represent the missing heritability of ASD (**Tables S8-S12**). Table 1 presents the highest frequency ASD-SVs, and their relevant biological processes.

**Table 1.** Number of genes associated with the biological processes identified in our Gene Ontology analysis in the SFARI database (N=816 genes represented by the array) compared to the ASD-SV identified in this study. Details for each gene are provided in the supplementary table listed on the left.

| Biological category | Genes in SFARI | Genes with ASD-SV | Suppl Table |
| --- | --- | --- | --- |
| Dendritic spinogenesis | 17 | 97 | S9 |
| Glutamate signaling | 24 | 120 | S10 |
| Chromatin modification | 28 | 114 | S11 |
| Axon guidance/migration | 11 | 60 | S12 |

Of the 19 genes that are annotated with the GO BP term “positive regulation of **dendritic spine morphogenesis**” (GO:0061003), 8 of them contained high frequency ASD-SVs. For example, nearly one-fifth of ASD individuals carry an ASD-SV in the Kalirin gene (*KALRN*, rs2120789), which is a RhoGEF that has been associated with schizophrenia ^36^. Involvement of this gene in spinogenesis was confirmed by reports demonstrating its disruption in mice produces altered dendritic density ^37^. This enriched group also includes the *RELN* gene, which has been associated with ASD in more than 50 studies (SFARI), and also its associated receptor LRP8. Both genes harbor high frequency ASD-SVs and both are necessary for proper dendritic spine development ^38^. In addition to the group of eight genes returned by the GO analysis, we obtained from the literature a larger group of genes linked to dendritic spine morphogenesis (N=97, **Table 1, Table S8**) and supported by *in vitro* and *in vivo* work, many of which contain high frequency ASD-SVs. For example, the brain-specific Kelch-like protein 1 (*KLHL1*) has been shown to causes dendritic deficits in mice when mutated ^39^ and copy number increases of the Necdin (*NDN*) gene, which lies at the terminal portion of the 15q11-q13 region we identified with consecutive SV-NMI (above and **Fig. S2**) causes increased spine density and hyperactivity ^40^. Many others indirectly participate in the manipulation of the actin cytoskeleton by regulating Rho GTPases such as the genes encoding GTPase-activating proteins, *ARHGAP24, ARHGAP15*, and *ARHGAP32*, the last of which likely causes the ASD-like Jacobsen Syndrome ^41^.

#### Glutamate Signaling

Significant enrichment for the GO term “synaptic transmission, glutamatergic” (GO:0035249) highlights the involvement of **glutamate signaling** in ASD (**Fig 2** and **Table S9**). Glutamate receptors mediate excitatory synapse transmission in the brain and are grouped into five families (AMPAR, NMDAR, Kainate, Delta, and mGluR), all of which have been implicated in ASD ^42,43 44^ and in the ASD-like Kleefstra Syndrome ^45^. Of the 26 genes that encode subunits of these receptors, we find that 20 harbor an ASD-SV, many at high frequency (F**ig. 4** and **Table S10**).

Importantly, a metabotropic glutamate receptor, *GRM5* (mGluR5), initiates a cascade of events that are central to dendritic spine formation ^28^, strongly connecting the biological functions amongst our ASD-SVs. We find that 22% of ASD cases harbor an ASD-SV in *GRM5* (marked by rs1846476), which intersects and is therefore predicted to disrupt a FOXA1 binding site, suggesting that *GRM5* is dysregulated in ASD individuals that carry this SV. Indeed, this was found to be the case in the recent single-cell RNA-Seq study ^46^. We find that several high frequency ASD-SVs reside in glutamate receptor subunits that are necessary for the early development of the cerebellum and are directly involved in development of the network of Purkinje cells and Climbing Fibers that are critical for the cerebellar function: *GRM5* (22%), *GRID2* (35%), *GRIA4* (5%), and *GRIN3A* (18%) (***Glutamate Signaling*** in Supplementary Text and **Fig. 4**). Further support is provided by an ASD-SV in *GRIN2A* that overlaps an open chromatin region necessary for fetal telencephalon development (rs6497523; **Table S8** and **Table S10**) ^47^. Indeed, nearly all post-mortem examinations of ASD brains have found significant differences in the cerebellum compared to controls, including the loss of Purkinje cells, overall cerebellar enlargement early in development, and reduction in size by adulthood ^48–51^. Together with the dendritic spine morphogenesis genes, the disruption to glutamate signaling genes supports the hypothesis that ASD is likely a disorder centered around aberrant development of the cerebellum.

#### Axon Guidance

Finally, the enrichment for genes involved in **neuronal migration** buttresses our claim that these ASD-SV represent a substantial component of missing heritability and the genes we identify interact with each other again supporting the claim that the heterogeneity of ASD results from disruption of different genes that participate in the same biological process. Live brain scans as well as post-mortem studies of ASD cases have identified an altered neuronal connectome ^52^. The development of complex neural circuits requires the migration of axons over long distances to make the appropriate connections to their target cells. This process requires an axon guidance “cone” at the tip, which senses attractant or repulsive cues secreted by astrocytes and other cells that lie along the path. The axons turn based on the combination of the molecule secreted and the receptor(s) being expressed at the tip of the cone. Upon passing a secreting sentinel cell, the receptors at the tip are degraded and replaced with new receptors that will sense the next decision point in the pathway. Often the axon will make contacts with the cell it passes via contactin and contactin-associated proteins (CNTNs and CNTNAPs).

The majority of the axon-guidance related genes harboring ASD-SVs are either the receptors expressed at the cone of the migrating axon, or their partner ligand that is secreted by the cells at the choice point (***Axon guidance*** in Supplementary text, **Table S11**). For example, we identified frequent ASD-SVs in the Unc-5 Netrin Receptor C (*UNC5D*, rs4699836, 29% of cases), its cofactor DCC Netrin 1 Receptor (*DCC*, rs9304422, 28% of cases), and the ligand Netrin G1 (NTNG1, rs4915019 in 26% of cases), which has been associated with ASD ^53^ and ASD-like RETT Syndrome ^54^. Similarly, two Roundabout Guidance Receptors (ROBO1 and ROBO2, rs4856257 and rs687813, 18% and 19% of cases respectively) and their ligands, Slit Guidance Ligands (SLIT3, SLIT2, SLIT1; rs7664347, rs888783, rs2636809 in 13%, 23%, and 13% of cases, respectively) carry ASD-SVs. Expression of *ROBO1* and *ROBO2* are significantly downregulated in ASD ^55^ and SVs have been reported in *ROBO2* in ASD cases ^56^. Variants in both ROBO3 and SLIT2 fully co-segregate with sound-color synesthesia (stimulation of one sensory input provokes perception in another), which is often comorbid with ASD ^57^. The distribution of ASD-SVs amongst several members of the same biological pathway and their previous association with the disorder are clearly non-random and provide even further support for our hypothesis that the NMI approach is identifying SVs that have previously gone undetected and explain missing heritability of ASD.

### Validation of ASD-SV disruption of glutamate signaling with RNA-seq data

Consistent with our observation that most of the ASD-SV reside in non-coding genomic and therefore likely regulatory space, we found them to be significantly enriched in 513 differentially expressed genes (DEGs) reported in post-mortem brain tissue from ASD cases and controls ^46^ that were completely independent from the populations from which we obtained the SNPs to identify the SVs. In this case, more than 70% of the DEGs (364 genes) harbor an ASD-SV, which is significantly greater than expected by chance (chi-square test, p < 3.0 × 10^−60^; **Fig. 3d, Table S5**). One of the most frequent ASD-SVs resides in the gene *GRIK2*, which encodes the GluK2 subunit of the kainate receptor (KAR, 35% of cases; **Fig. 4, Table S10**) previously associated with ASD ^60^ and, in line with convergence of ASD-SV to a few biological processes, is central to dendritic spine formation ^61^. The SNP (rs2051449) that marks this ASD-SV offers an opportunity to delve deeper into the genetic disruption linked to ASD because the NMI approach provides kilobase-resolution as to the locale of the SV. In this case, the ASD-SV overlaps a DNAse I hypersensitive site with a known CNV ^62^ adjacent to exon 12 that binds an RNA-splicing complex (**Fig. 5a**). An SV at this site is therefore predicted to disrupt proper splicing of exon 12. Exon 12 codes for a portion of the glutamate binding pocket and therefore the loss of this exon would significantly disrupt glutamate signaling, especially as it is predicted to still be capable of assembling with other subunits via the preserved amino-terminal domains, which would result in a loss of function via a dominant negative mutation (**Fig. 5a** and ***Glutamate Signaling*** in **Supplementary file**).

**Fig. 5.**
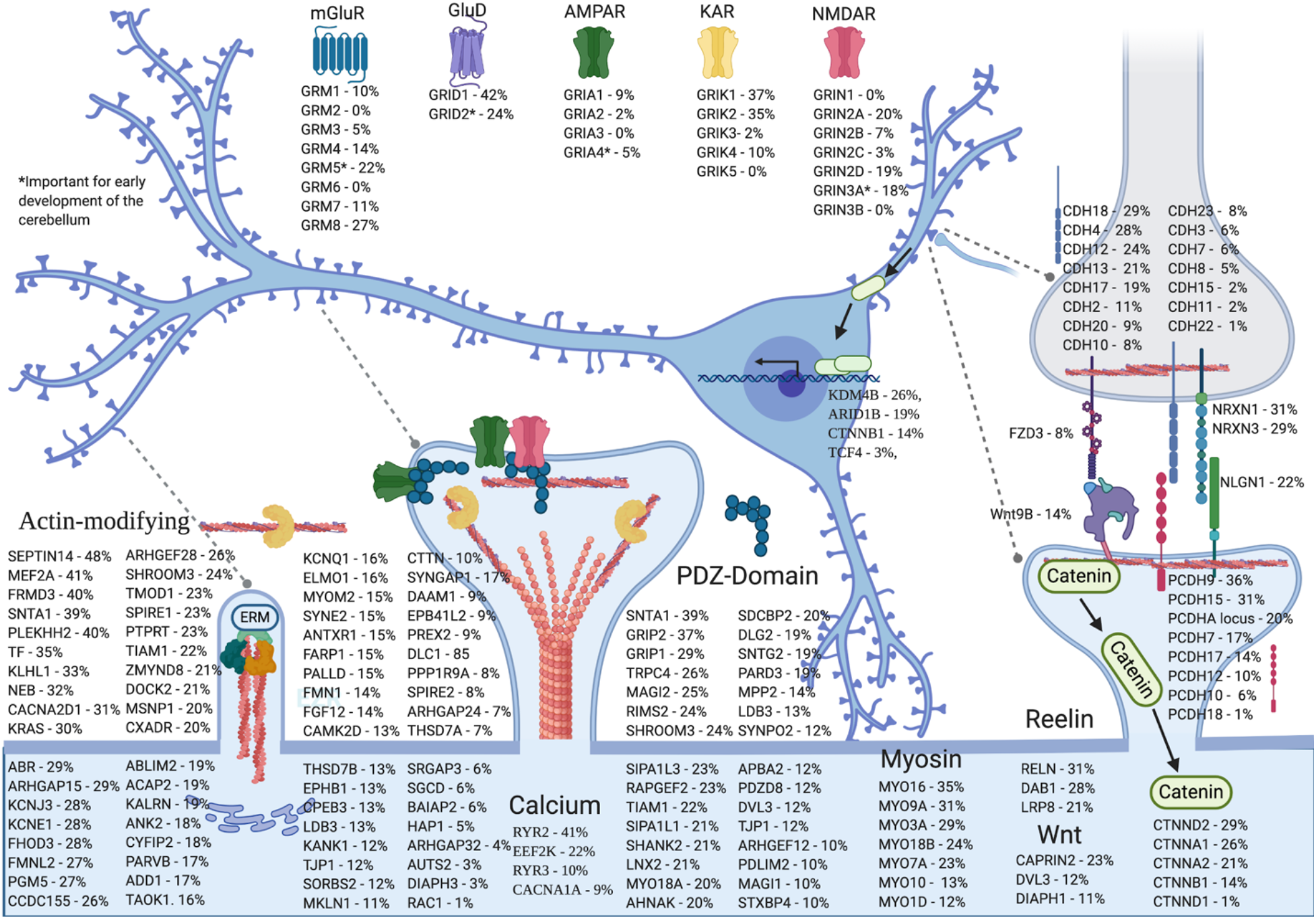
Dendritic morphogenesis and ASD-SV frequency. The formation of dendritic spines (lower blue-shaded processes) involves proteins of diverse functions that generate synapses with axons (upper gray-shaded process), many of which our method indicates are disrupted by SVs in individuals with ASD. The most numerous are those that directly manipulate the actin cytoskeleton to form the spine (N=97 genes including *MSNP1* from the transmission disequilibrium test). GRM5, NMDA, and AMPA receptors mediate calcium release and phosphorylation of moesin (MSN), which is a major regulatory point of spine formation and is itself regulated by *MSNP1* (**Fig. 4**). The glutamate signaling pathway is activated by Wnt/β-catenin signaling (green ovals) via TCF4 and the H3K9me3 lysine demethylase KDM4B ^58^ and is repressed by ARID1B ^59^. This effectively links three of the four processes identified in the GO enrichment analysis. Many of the most frequently affected glutamate receptor subunits are involved in the early development of the cerebellum (see Section 2.6 and **Supplementary file**). Mean frequency of ASD-SVfor the two ASD studies is provided for each gene (individuals with NMI pattern/total individuals studied).

We re-analyzed the data from Velmeshev et al. at the exon level, which revealed a roughly 50% reduction in transcripts within exon 12 in 10/13 ASD samples but in only one of the controls (**Fig. 5b**), thus providing stronger evidence of disruption of glutamate signaling in ASD due to an SV adjacent to exon 12. In order to further interrogate the role of *GRIK2* in ASD and find potential links to other ASD-SVs, we first performed a differential gene expression analysis of the nine controls that retained *GRIK2* exon 12 versus the ten ASD samples that showed reduced transcripts within *GRIK2* exon 12. This identified 2,685 significantly differentially expressed genes (FDR < 0.05; **Fig. 5c, Table S12**). Similarly, we split the AGPC data set into two sub-groups: those with and those without the SV at SNP rs2051449, based on a plot of the intensity values (**Fig. 5c**). We identified 15 ASD-SVs that had significantly differentially observed frequencies (DOSV) between the two groups (**Table S1**3). Two of those ASD-SVs were in the *PTPRD* gene, whose mRNA was also found to be differentially expressed in the post-mortem prefrontal cortex in ASD individuals ^46^. Furthermore, both *PTPRD* and *GRIK2* were previously identified in a GWAS as strongly associated with obsessive-compulsive disorder, which is highly comorbid with ASD ^22^. A plot of the expression of *GRIK2* and *PTPRD* reveals that they are co-regulated in controls but not in ASD individuals (**Fig. 5c**). As is the case with *GRIK2, PTPRD* regulates dendritic spine formation, further supporting the role of disruption of this process by SVs as core to ASD. Notably, the most frequent ASD-SV in *PTPRD* (rs7026388) lies within an exon, suggesting it disrupts the protein. It is highly noteworthy that most ASD individuals carry an ASD-SV either in *PTPRD or* in *GRIK2*, again consistent with the proposed molecular heterogeneity of the disorder, i.e. disruption of only one of those genes can result in ASD as they affect the same biological process.

**Fig. 5.**
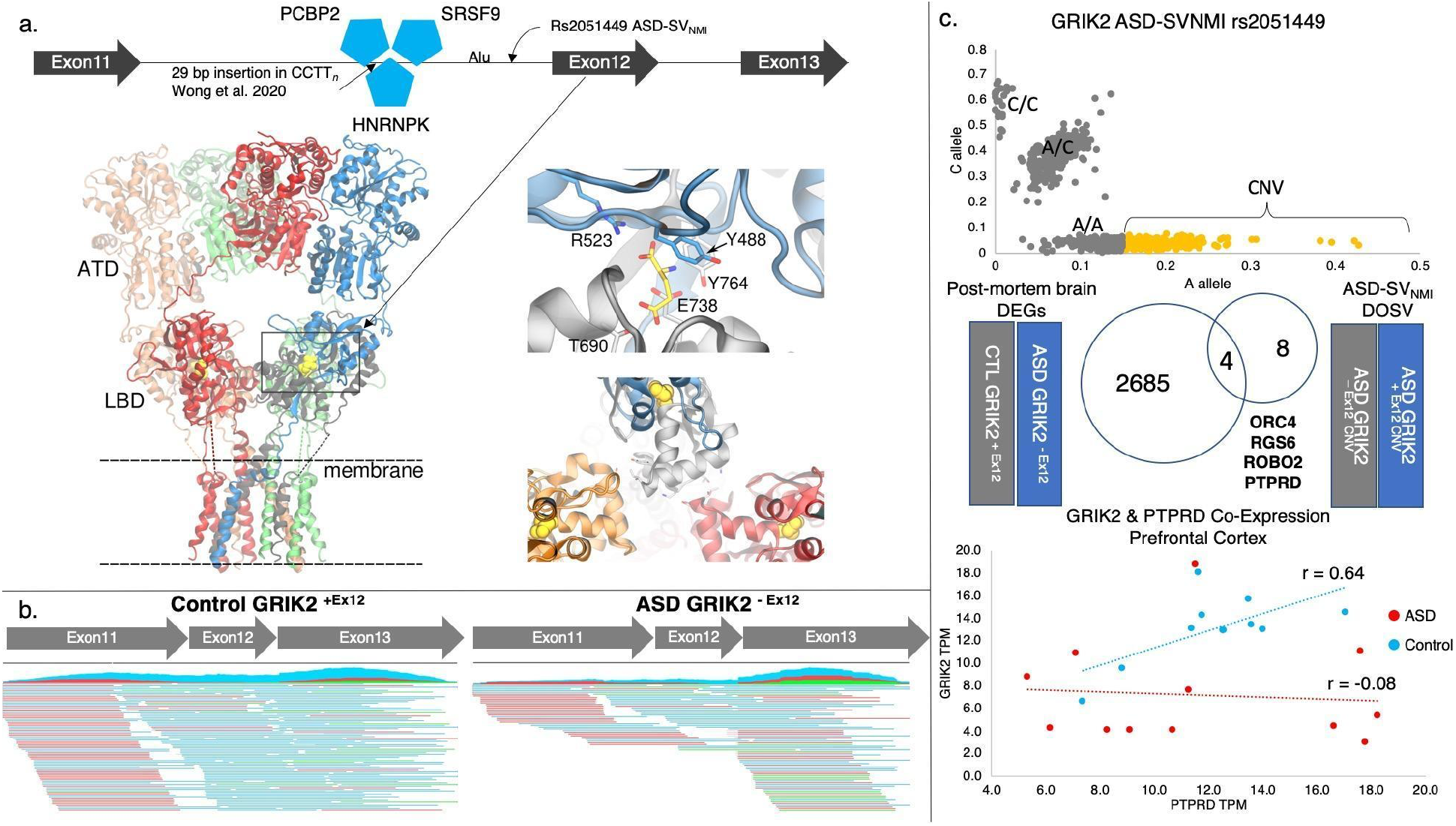
ASD-SV impair glutamate signaling associated with disruption of the GluK2 (encoded by *GRIK2)* (**a**) The ASD-SV at SNP rs2051449 is predicted to disrupt a known splice site adjacent to exon 12 bound by PCBP2, SRSF9, NHRNKP identified from the ENCODE project. A recent analysis of SVs from 388 individuals of diverse ancestry identified a 29 base pair insertion at a CCTT_*n*_ repeat near this site ^62^. Exon 12 is important for glutamate binding. Each subunit of the tetrameric GluK2 (*GRIK2* gene) is composed of an amino-terminal domain (ATD), a ligand binding domain (LBD) and a transmembrane domain. The subunits are distinguished by color (orange, green, red, and blue) and amino acids coded by exon 12 are illustrated in one subunit, in grey (left structure). The cryo-EM structure of the complex from *Rattus norvegicus*, which is 99% identical to the KAR from *Homo sapiens*, was used here (PDB 5KUF). Main amino acid residues in contact with the glutamate ligand are depicted (in yellow, magnified top right). T690, E738 and Y764 are absent due to missing exon 12 in *GRIK2* (PDB 4UQQ was used to represent the binding site with glutamate). The region encoded by exon 12 interacts with adjacent LBDs (magnified bottom right) and is critical to the functional dynamics of the tetrameric GluK2 ^63^.. (**b**) Mapping of RNA-seq data from post mortem brain tissue reveals 10 of 13 ASD individuals display loss of exon 12 whereas only 1 of 10 controls do (**c**) Plotting of the intensity signals for rs2051449 indicate a likely copy number gain at the site. Partitioning of 26,524 SVs into those with and without a CNV at rs2051449 identified 12 differentially observed coding ASD-SV (FDR < 0.05, DOSV, two in the same gene, *PTPRD*). Four genes intersected with differentially expressed genes (DEGs) from post mortem brain tissue from (b). *PTPRD* and *GRIK2* expression levels are significantly correlated in prefrontal cortex from control individuals (0.65, *p* < 0.03) but not those with ASD (−0.08, *p* < 0.79), further supporting the role of the disruption of these genes as a core component of ASD.

### ASD-SVs provide an important marker set for association with phenotype

We performed logistic association using a set of presence/absence markers encoded for ASD-SVs located within genes (*see* Data Description and Methods) and verbal/non-verbal phenotype data. The test identified two significant loci, *ACMSD* and *MTHFD2P1*, after a conservative Bonferroni correction (p < 5 × 10^−6^, **Fig. 6a**). ACMSD is an important enzyme in the tryptophan/kynurenine pathway, and is responsible for producing the neuroprotective picolinic acid from quinolinic acid substrate (**Fig. 6b**). Both the product and substrate have been linked to schizophrenia, Tourette’s syndrome, epilepsy, depression, suicide, and importantly, ASD ^64–67^. Here, the significant ASD-SV occurs at a SNP (rs12471304) 1 kb from a FOS transcription factor binding site that has been reported to regulate the *ACMSD* gene in the Open Regulatory Annotation database (OREG1613578) ^68^.

**Fig. 6.**
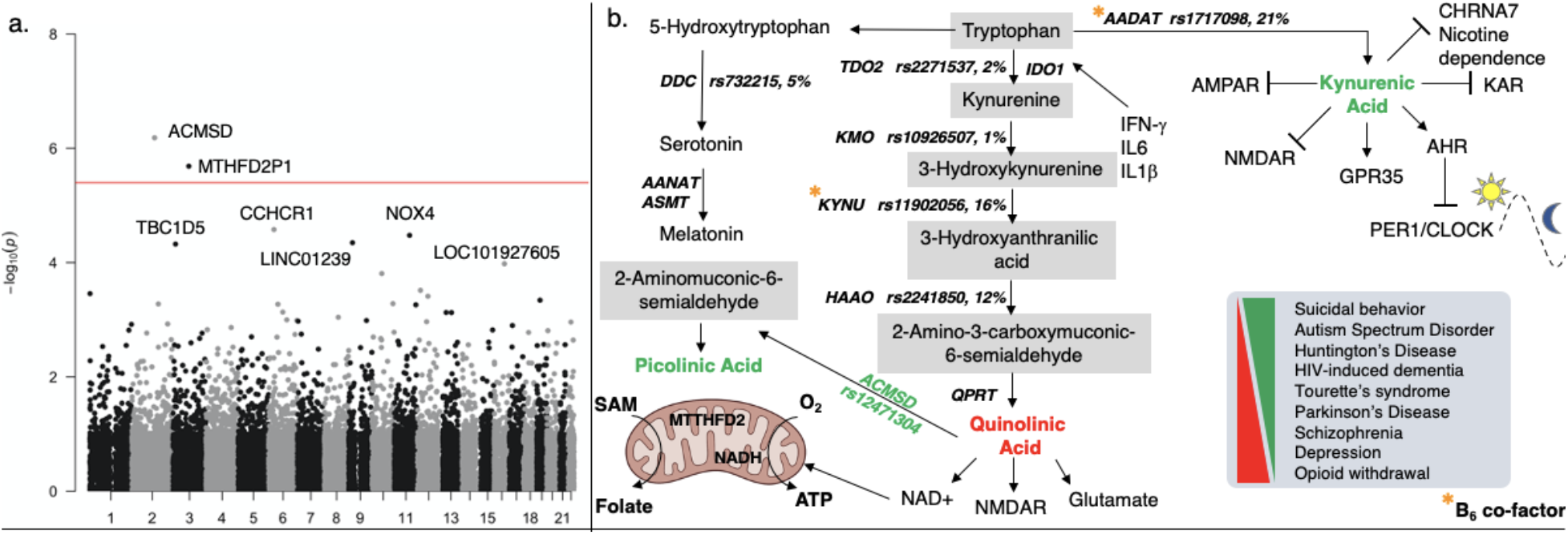
Association testing of ASD phenotypes using ASD-SV markers. (**a**) Manhattan plot of association testing of verbal vs. non-verbal phenotype using presence/absence markers of ASD-SVs at 10,108 loci found two significant ASD-SVs after Bonferroni correction (red line). (**b**) The most significant association resides in a FOS transcription factor binding site that regulates the *ACMSD* gene, which codes for a key enzyme in the kynurenic acid pathway. Altered levels of quinolinic acid and picolinic acid of this tryptophan catabolic pathway have been associated with several neuropsychiatric disorders including ASD ^66^, and a SNP in this gene has been linked to suicidal behavior ^65^. The metabolites kynurenic acid and quinolinic acid in this pathway inhibit glutamate signaling via numerous receptor types, one of which (NMDAR) is a therapeutic target for the treatment of ASD ^72^.

In addition to picolinic acid and quinolinic acid, tryptophan can also undergo catabolism to kynurenic acid through action of the enzyme aminoadipate aminotransferase (AADAT), which inhibits NMDA, Kainate, and AMPA receptors. A report of altered plasma levels of kynurenic acid and tryptophan in ASD cases compared to controls and correlation with disorder severity further supports our findings here ^67^. As is the case with picolinic acid, kynurenic acid appears to be neuroprotective ^69,70^ (**Fig. 6b**). Notably, an ASD-SV at rs1717098 in *AADAT* is found in more than 20% of individuals in both the MIAMI and AGPC studies. The SV overlaps a regulatory site for *AADAT*, and a CNV in ASD cases has been reported in this gene ^71^. As with the biological pathways identified by our GO tests, our association test between verbal and non-verbal cases with only genomic regions harboring ASD-SVs pinpoint a specific pathway with multiple affected genes that has already been strongly associated with the disorder in previous studies.

### Clustering of ASD-SVs reveals the genetic heterogeneity of autism

By using an explainable artificial intelligence (X-AI) approach, we demonstrate that we can use the ASD-SVs to dissect the heterogeneity that has plagued past studies, providing further support that these genomic variants represent a large component of the missing heritability of ASD. Using hierarchical clustering we were able to delineate several distinct sub-clusters of the AGCP ASD cases (**Fig. 7a**). Then, by using an iterative Random Forest classifier ^73^, we identified the genes whose SV variation across the ASD cases most defined each cluster (**Fig. 7b**). This provides invaluable information for follow-up studies. For example, an ASD-SV in the *CTNNA2* gene defines cluster number 1 and is associated with the startle response ^74^, whereas the *CACNA2D1* gene, which defines cluster 3, is associated with Long QT cardiac arrhythmias ^75^. These NMI variants could be tested for association with distinct ASD phenotypes.

**Fig. 7.**
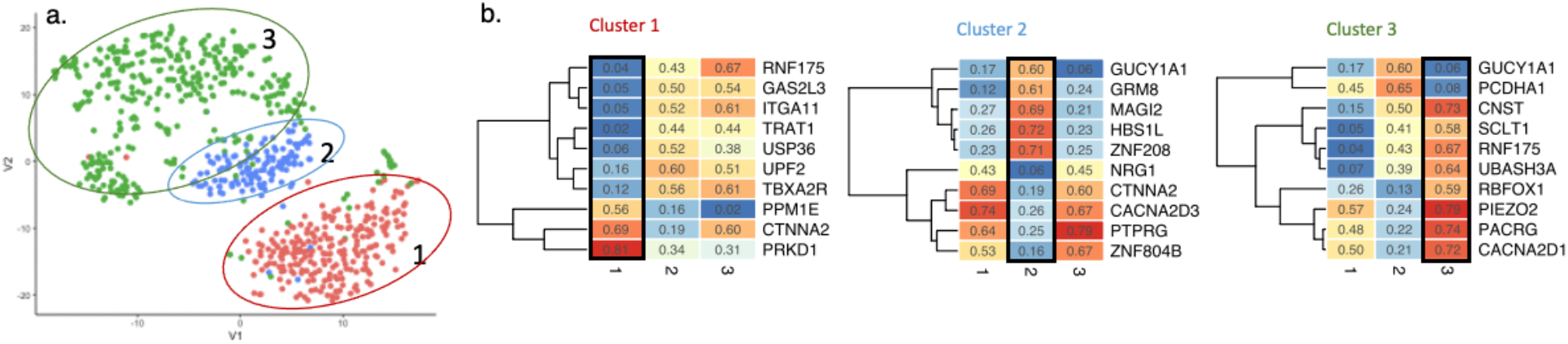
Identification of ASD subgroups from GWAS. (**a**) tSNE plot colored according to hierarchical clustering of genic ASD-SVs shows three subgroups of ASD individuals from the AGPC study. (**b**) ASD clusters can be explained by the most important genes containing ASD-SV according to iterative Random Forest classifiers. The top 10 genes (based on RF importance score) for a cluster are shown in a heatmap where cells are colored according to the frequency of their resident ASD-SV (blue = low frequency, red = high frequency) and the contrast with the other two clusters is evident in each heatmap. Frequency values are shown in the cells.

## Discussion and Concluding remarks

Using simple NMI as a surrogate for SV detection, we uncovered several lines of evidence that recapitulate the well-described ASD phenotype and molecular pathways and provide further insight into how they are integrated. First, we confirmed the most strongly associated locus from one of the source studies (*MSNP1*) but with much greater statistical significance. Second, because we were able to substantially increase signal over noise compared to previous work, a biological enrichment test confirmed the widely accepted view that dendritic spinogenesis is a core biological process that underlies ASD and identified a myriad of SVs that may affect it. This is encouraging for the current focus on the development of pharmaceutical interventions that target the p38 pathway because it appears to be a core regulator of dendritic spine development and neural plasticity ^76–78^. Importantly, these ASD-specific SVs define links between what have previously been regarded as disconnected biological processes. Due to the numbers and specific genes we identify that harbor ASD-SV, we show that glutamate signaling, neuronal migration, synaptic transmission, and chromatin modification are also critical for dendritic spinogenesis and axon migration during brain development. Third, because the NMI method pinpoints the location of the SV, we were able to test and validate the molecular mechanistic hypothesis that an SV in the *GRIK2* gene disrupts splicing of an exon coding for the GluK2 glutamate-binding pocket. Further analysis of this RNA-seq data from unrelated individuals independently confirms a previous report of the association of variants in *MSNP1, GRIK2*, and *PTPRD* in neurodevelopmental disorders.

Critically, our genome wide association analysis with the non-verbal phenotype using only ASD-SV supports the hypothesis that the missing heritability of ASD resides in SVs. It also implicates the kynurenine pathway in the disorder, which lies at the nexus of numerous ASD-associated traits including neuroinflammation, sleep disorder, gastrointestinal abnormalities, and altered circadian rhythms, as well as supports the major involvement of glutamate signaling imbalance in ASD.

Our results indicate that the failure of standard methods such as GWAS to detect loci that are strongly associated with ASD is not due to any shortcomings of the method, but rather the inability of current genotyping methods to provide accurate information for testing. Until either sequencing or variant calling methods improve, we have shown here that much of this important genomic variation can be captured with simple NMI signals on genotyping arrays. Re-analyses of existing pedigrees with SNP genotypes would be a logical first step. In the absence of family-based information, GWAS approaches could still be used without standard filters, such as HWE tests, to obtain at least some of the signals we identify with NMI.

Lastly, this study emphasizes the importance of re-analyzing existing data sets with new tools and approaches. With the novel database of SVs created here, we were able to characterize, at the molecular level, a set of ASD individuals that are likely to be non-verbal. We developed this database using data that is customarily excluded from analysis. It is therefore very likely that such previously undetected SVs are the key “missing heritability” needed to explain the high heritability of ASD (*h*^*2*^ = 0.80) and other human diseases and phenotypes. We predict this approach will rapidly advance the knowledge of the genetic basis of many health conditions of societal importance.

## Materials and Methods

### Samples and Quality Control

We obtained array-based genotypes from ASD cases and their parents from the database of Genotypes and Phenotypes (dbGaP). For SV discovery we used a dataset from an ASD study from the University of Miami ^15^ consisting of 1,177 individuals that represent 381 families genotyped at 1,048,847 nuclear SNP loci (dbGAP accession phs000436.v1.p1). We labeled this dataset as MIAMI. For validation we used data from a second study ^14^, which was produced by the Autism Genomic Project Consortium (AGPC), and consists of 4,168 individuals representing 1,385 families genotyped at 1,072,657 nuclear loci (dbGAP accession phs000267.v5.p2). We labeled this dataset as AGPC. Data were handled in accordance with the rules established by the National Institutes of Health. Potentially erroneous SNPs were removed by excluding all those with a quality score of less than 0.75, and we performed a kinship analysis to ensure there was no overlap between individuals in MIAMI and AGPC (**Methods in Supplementary file)**.

### Non-Mendelian Inheritance (NMI) Detection and re-genotyping

We used the program PLINK v1.9 ^79^ with the 890,539 autosomal SNPs that remained after QC filtering to identify loci that did not conform to Mendelian inheritance and therefore represent likely SV. We did not include SNPs on the X chromosome because NMI cannot be determined on the X in males due to hemizygosity. In most cases of NMI that we observed, the Mendelian expectation was that the child should be heterozygous at a site but instead displayed homozygosity (Fig 2, Fig S1). There are a considerable number of cases where an SV may exist and be causing erroneous genotype calls, but PLINK does not detect NMI at that site because all three members of the trio show the same homozygous genotype (e.g., all are A/A). However, if a number of other trios at that site do have clearly detectable NMI patterns, then we can leverage their genotyping signal intensities to find SVs in individuals not called by PLINK (Fig 1b, center). If an individual’s genotype intensity co-located on a signal plot with those with NMI, then we marked this as “*suspected NMI*” and can infer the presence of an SV in that individual. Once individuals were marked as *NMI* or *suspected NMI* at a site, we re-genotyped them according to signal intensity plot positions (Fig 1b, right).

The *mendel* function in PLINK outputs codes that can be directly translated into paternal or maternal errors. In addition, some NMI trio genotype combinations are ignored by PLINK, so these were scored manually and combined with the scored sites into a single matrix of genotypes for each of MIAMI and AGPC. For example, we scored scenarios where genotypes were child = “A/A”, father = “A/A”, and mother = “-/-”, assigning it as a maternal SV. Paternal SV was assigned when the genotype is missing for the father but present in the mother. In this matrix the sites represent putative SVs of indeterminate length, though an upper bound of length can be derived by observing the basepair distance to the next normal mendelian site on the array. The NMI genotyping workflow can be seen in Fig 2. We used the smaller MIAMI data set (N=381 families) for SV discovery and the large AGPC data set (N=1136 families) for validation (**Supplementary file** for a detailed description).

### Filtering NMI SVs

Our goal was to reduce the initial set of NMI sites to a set of reliable ASD-specific SVs that are most likely to represent the core of the missing heritability of ASD. Fig. 2 provides an overview of the workflow described below.

First we applied filters to remove potential false positive SV genotypes. Rarer SVs are more likely to be due to error than common SVs, so we removed all SVs with frequency less than 2% in the discovery population (MIAMI). We chose 2% because this is the estimated frequency of ASD in humans. It is also an extremely conservative filter given that the technical error rate for the Illumina array used in this study was estimated to be less than 0.05%. A potential cause of a false positive genotype for an array SNP is the presence of other SNPs in the immediate genomic region of the probe for that SNP.

Therefore, we also removed any SV whose probe overlapped another SNP (according to dbSNP153) with a MAF > 0.02 in the 1KGP EUR population. Finally, SVs that are found in only the discovery dataset are more likely to be false positives, so we intersected the NMI SVs discovered in the MIAMI population with those in the AGPC validation population and removed any which did not appear in both. The resulting set of higher confidence SVs was labeled as **NMI-SV**.

Next we reduced the NMI-SV set to a subset of novel ASD-specific SVs by removing those whose genotyping probe intersected with previously identified SV intervals with MAF > 0.02 in one or more non-ASD-specific sources (**Methods in Supplementary file**). Sources included the 1000 Genome Project hg38, a long-read sequencing scan from the same population ^80^, 433,371 SVs identified from 14,891 diverse genomes ^81^, and a recent report of 107,590 SVs (most of them novel) from genome-scale resolved haplotypes ^11^. To be conservative, we removed NMI-SVs in this manner even if they resided in a gene that had previously been identified as ASD-related (*see NRXN3*, Fig 1). The NMI-SVs that appeared in both ASD study populations and passed through all filters were labeled as **ASD-SV**s. Finally, we reasoned that the core biological pathways in ASD would be represented by the most frequent ASD-SVs, so we defined a core set of ASD-SVs found in both study populations at greater than 15% frequency.

### Detecting large SVs

To identify large SV (runs of NMI in each individual), we calculated a running sum on position-sorted NMI with a window size of 5 SNPs and calculated the probability of obtaining 5 sequential NMI SNPs on arrays that were randomized, i.e., SNPs that are adjacent on a chromosome are spread randomly across each array. The binomial probability of obtaining 5 successes (k) in 5 trials (n) with a probability of success of 0.36% (p) is 6 × 10^−13^.

### Gene Enrichment

The set of genes harboring NMI-SVs were subjected to enrichment tests to determine if they were functionally non-random. We used a chi-square test to see if these genes were enriched for ASD-susceptibility protein-coding genes listed in both SFARI (sfari.org/resource/sfari-gene/ in April 2021)) and AutDB (autism.mindspec.org/ in April 2021) databases.

The set of genes harboring core ASD-SVs (those with freq > 15% in both populations) were assessed for enrichment for Gene Ontology biological process (GO BP) terms (http://geneontology.org/) with a false discovery rate (FDR) < 0.05. Additionally, we performed a permutation test by computing GO enrichment on 100 randomly sampled sets of 1,106 genes from a list of all genes that overlapped SVs identified from fully-resolved genome wide-haplotypes in the 1000 Genome population (N=5,810 protein coding genes, **Table S7b**) ^11^. Functional analyses for specific genes were taken from GeneCard Human Gene Database. ToppGene (https://toppgene.cchmc.org/) was used for the disease associated enrichment test of the core ASD-SV genes.

### Transmission Disequilibrium

We tested for transmission disequilibrium for SVs in loci where there were no known SVs detected in the 1000 Genome EUR population. To do this, we used a χ-square test in which the expected number of SVs for males and females was calculated based on the expected frequency of 50%. We used a Benjamini and Hochberg FDR correction for multiple tests using the *p*.*adjust* function in R and identified four loci that were significantly greater for paternal inheritance and nine that were greater for maternal inheritance (**Table S6a**; FDR < 0.05). The top paternal hit (rs11739167, 17.6 fold greater than expected, *punadj* < 5.8 × 10^−14^) is centered on the pseudogene for membrane-organizing extension spike protein (*MSNP1*) (**Supplementary file**).

### Gene Expression Analysis for GRIK2

We downloaded RNA-seq FASTQ files for 13 ASD cases and 10 controls from Velmeshev et al. ^46^ from bulk prefrontal cortex listed in project PRJNA434002 in the sequence read archive (SRA) at NCBI. Reads were trimmed with CLC Genomics Workbench (version 20.0.4) then mapped to the human transcriptome GRCh38_latest_rna.fa with the following modifications: (1) predicted mRNA sequences were removed (those with the prefix “XM”), (2) all *GRIK2* transcripts were removed and replaced with a single transcript containing only exons 11, 12, and 13. This was done to reduce bias from reads mapping to UTRs and to focus on potential loss of exon 12 because this is the exon adjacent to the ASD-SV and predicted to be lost from aberrant splicing. Mapping parameters were set to 0.95 for both length fraction and similarity fraction to reduce mis-mapping of reads from closely related genes (e.g., *GRIK1* and *GRIK2*). The CLC Genomics tool Differential Expression for RNA-Seq was used with TMM normalization to control for library sizes. This tool assumes a negative binomial distribution for read counts similar to EdgeR and DESeq. Correlation between *PTPRD* and *GRIK2* expression was determined with a Pearson correlation test in the R package Hmisc. Significance was determined with an FDR correction < 0.05.

### Association testing for verbal/non-verbal forms of ASD

In order to perform a Genome Wide Association Study using ASD-SVs we first collapsed all ASD-SV sites within a gene’s boundaries (according to RefSeq) to a single presence/absence marker. If at least one of the ASD-SVs sites in a gene was present for an individual, then an ASD-SV was considered as present in that gene, even if the other sites were absent. Those sites that were not assigned to a gene by RefSeq were annotated with their rsID, and loci found at less than 5% frequency were removed, leaving 10,108 presence/absence markers for further analyses. We performed a logistic regression in PLINK and used the first two components of a PCA generated from 42,761 neutral SNPs as covariates to account for substructure of the ASD population (**Methods in Supplementary file**). The verbal (control) and non-verbal (case) phenotypes were extracted from the meta data included with the dbGAP project.

### Classification of ASD subtypes based on genic ASD-SVs

By collapsing core ASD-SVs within gene boundaries, we obtained presence/absence markers in the larger AGPC population for 1106 genes with frequency > 15%. Sub-structure within the presence/absence matrix was visualised in two dimensions using tSNE in R. We then applied hierarchical clustering using hclust with Bray-curtis distance and ward.D2 method in R, and selected clearly defined clusters as putative subtypes of ASD. In order to determine which genes have presence/absence patterns that define these subtypes, we used a custom R implementation of iterative Random Forest (iRF) machine learning ^73^ to classify the cluster labels. To do so, we set the labels for individuals in a single cluster to 1, and the rest to 0. The presence/absence for each gene was set to 0/1 and all genes were used as features in the iRF model, which performs an iterative feature selection. This process was repeated for each of the clusters, resulting in a final random forest for each cluster. The top 10 most important genes for each cluster were extracted based on their Gini importance scores provided by the Ranger v0.12 R package ^82^.

## Data Availability

Data were obtained through the NIH database of genotypes and phenotypes and handled in accordance with their guidelines.

## Data Availability

All original genotype data was obtained from the Database of Genotype and Phenotypes project numbers phs000267.v5.p2 and phs000436.v1.p1. Results from our analyses are provided as Supplementary Tables uploaded with the manuscript.

## Credit Contributions

David Kainer (Conceptualization, Formal Analysis, Investigation, Methodology, Validation, Visualization, Validation, Writing - original draft, Writing - review & editing), Alan Templeton (Conceptualization, Supervision, Writing - original draft, Writing - review & editing), Erica Prates (Formal Analysis, Investigation, Visualization, Writing - original draft, Writing - review & editing) Euan Allan (Formal Analysis, Investigation, Writing - original draft, Writing - review & editing), Sharlee Climer (Conceptualization, Supervision, Writing - original draft, Writing - review & editing), Daniel Jacobson (Funding acquisition, Methodology, Writing - original draft, Writing - review & editing), Michael Garvin (Funding acquisition, Conceptualization, Data Curation, Formal Analysis, Supervision, Investigation, Methodology, Software, Visualization, Validation, Writing - original draft, Writing - review & editing).

## Additional files

### Supplementary files

- Supplementary File 1. Supplementary Methods, Fig.s S1-S2, Supplementary Text
- Supplementary File 2. Excel Tables S1-S13

