## Supplementary file for "Structural Variants Are a Major Component of the Missing Heritability of Autism Spectrum Disorder"

**Supplementary Methods**

*Sample processing*

Potentially erroneous SNPs were removed by excluding all assays with a quality score of less than 0.75. One family was removed from the Miami data set and two from AGPC due to poor data quality and 248 families were removed from AGPC because they did not have a quality score listed with the genotypes or were not part of a trio (i.e., those missing one or both parents). In order to ensure we were analyzing two independent sets of parent-child trios, we performed a kinship analysis on all of the individuals from the 380 families from the University of Miami study and the 1,136 families from the AGPC study. We randomly chose 50,000 SNPs that conformed to Hardy-Weinberg-Equilibrium (HWE) and Mendelian inheritance, and had a minor allele frequency (MAF) of greater than 0.05. We also pruned SNPs that had an LD > 0.20 using the default step and window size on PLINK 1.9. We then removed any SNPs in which alleles were INDELs, A/T or G/C pairs, or were found on the pseudoautosomal regions of the sex chromosomes, leaving 48,478 SNPs for further analysis. We used the KING function in PLINK2 to estimate kinship. Kinship estimates within families were as expected. We identified a single female that was listed in two different trios within the AGPC study, which was consistent with the metadata as she was the mother in different trios (different fathers). No individuals were identified among trios that would indicate overlap of the Miami and AGPC data sets. In order to identify potential substructure of the ASD population, after excluding all loci that demonstrated NMI as potential SVs, we randomly chose 50,000 SNPs from the remaining assays. After intersecting with the 1000 Genome population and excluding those with MAF < 0.05, we retained 42,761 for the PCA performed in PLINK.

*High-confidence SVs*

Using the Miami and AGPC datasets, we performed an NMI test in PLINK on both sets of data, which flagged 101,032 SNPs having at least one family with NMI in one of the data sets. We then manually scored these 101,032 sites for NMI in further families that PLINK did not flag and estimated the frequencies within each population (Fig. S1). All SVs found at a frequency of less than 2% in the Miami set were removed, leaving 61,703 as our discovery panel. We chose 2% because this is the estimated frequency of ASD in the human population but also an extremely conservative filter given that the technical error rate for the Illumina array used in this study was estimated to be less than 0.05%. The 2% NMI rate corresponds to 7 individuals from the 380 families. The binomial probability of having a SNP assay fail 7 times in 380 trials given the technical error rate of 0.05% is 1.4 x 10^-9^, where *p*=0.05, *n*=380, and *k*=7. It should be noted that the quality control of the Illumina bead arrays releases assays that display the technical error rate of 0.05% or less, i.e., it does not account for error rate due to the samples being analyzed. Therefore, by definition, the error rate of 2% is conservative given that it is 40 times higher than technical background error.

Of this set, 90% (55,767) were found in at least one individual in the AGPC population. Next, we used a Pearson correlation test with the rcorr function in the package *Hmisc* in the R programming environment and calculated a significant correlation between NMI SNPs in the discovery and validation data sets of 0.75 (p < 0.0001). To identify large SV (runs of NMI in each individual), we calculated a running sum on position-sorted NMI with a window size of 5 and calculated the probability of obtaining 5 sequential NMI SNPs on arrays that were randomized, i.e., SNPs that are adjacent on a chromosome are spread randomly across each array. There were a total of 338,404,820 genotyping assays in the Miami data set (380 families x 890,539 SNPs used). Of these, 1,227,413 displayed an NMI pattern, or 0.36% of total genotyping assays across the 380 arrays. The binomial probability of obtaining 5 successes (*k*) in 5 trials (*n*) with a probability of success of 0.36% (*p*) is 6 x 10^-13^.

*ASD-associated CNV*

The AutDB CNV database was filtered for all cases with an ASD diagnosis for which there were genomic locations identified for the hg38 version of the human genome and overlapped at least one SNP from the Illumina Array and a genomic feature (N=22,233 cases, Table S4). We then intersected a BED file of these CNV with the ASD-SV to identify any that overlapped with the array. Because we can already identify large CNV using runs of NMI SNPs, here we wanted to focus on short CNV and therefore only included those that overlapped either one or two SNPs. CNV that overlapped a SNP with a minor allele frequency (MAF) of less than 0.001 were removed because they could not be discoverable with NMI. This left 2,270 CNV as a truth set. Of these, we identified 1,902 with NMI (84%). Although the NMI proved to be a robust method to detect known CNV, we wished to determine if lower allele frequencies of the SNPs that overlapped CNV could explain the inability to detect the remaining 16%. We compared the MAF of the 1037 SNPs that overlapped the CNV that were successfully detected with NMI to the 207 SNPs that overlapped CNV yet were unable to detect them by NMI. Those SNPs that failed to detect CNV demonstrated a significantly lower MAF compared to those that succeeded (p < 2.2 x 10^-16^, one-sided Wilcoxon rank sum test).

*Differential Observed SV with GRIK2 ASD-SV at rs2051449*

In order to determine if any ASD-SV were co-segregating with the one identified at rs2051449 in *GRIK2*, we first plotted the genotypes using the original Illumina array intensity values as was done for the individuals at the *NRXN3* SV_NMI_. In this case, the pattern suggested that there were copy number *gains* linked to the A allele and we therefore selected from the 1137 AGPC individuals the subset of those whose intensity value at the A allele was greater than those found in any of the heterozygotes. This is a conservative estimate of those with a gain because heterozygotes harbor only a single A allele and therefore intensities will be lower than homozygotes. We calculated the expected number of each ASD-SV based on the overall frequency in the AGPC population (381 with and 756 without the ASD-SV at rs2051449) and tested for significance with a *Chi*-squared test. Because this test is unreliable at low numbers, we only included ASD-SV that were found in at least 20 individuals. Of these 26,524 ASD-SV, 15 were found to be differentially observed (FDR < 0.05). FDR was calculated using the p.adjust function in R with the Benjamini & Hochberg method. All significantly different ASD-SV were found at lower than expected numbers and two were identified in the same gene, *PTPRD*.

*Association tests for verbal/non-verbal forms of ASD*

In order to perform a Genome Wide Association Study using ASD-SV we first collapsed all sites within a gene’s boundaries (according to RefSeq) to a single locus. If at least one of the ASD-SV sites in a gene was present for an individual, then an ASD-SV was considered as present in that gene, even if the other sites were absent. Those sites that were not assigned to a gene by RefSeq were annotated with their rsID, and loci found at less than 5% frequency were removed, leaving 10,108 loci for further analyses. We performed a logistic association in PLINK and used the first two components of the PCA generated from 42,761 neutral SNPs (see *1.1 Sample processing*) as covariates to account for substructure of the ASD population. The verbal (control) and non-verbal (case) phenotypes were extracted from the meta data included with the dbGAP project.

*Classification of ASD subtypes based on genic SVs*

By collapsing ASD-SV sites within gene boundaries, we obtained presence/absence markers in the AGPC population for 1106 genes with frequency > 15%. Sub-structure within the presence/absence matrix was visualised in two dimensions using tSNE in R (Fig 8a). We then applied hierarchical clustering using *hclust* with Bray-curtis distance and ward.D2 method in R, and selected 3 clearly defined clusters as putative subtypes of ASD. In order to determine which genes have presence/absence patterns that define these subtypes, we used a custom R implementation of iterative Random Forest (iRF) machine learning [[20]](https://paperpile.com/c/0BBqdN/L8RP) to classify the cluster labels. To do so, we set the labels for individuals in a single cluster to 1, and the rest to 0. The presence/absence for each gene was set to 0/1 and all genes were used as features in the iRF model, which performs an iterative feature selection. This process was repeated for each of the three clusters, resulting in three final random forests. The top 10 most important genes for each cluster were extracted based on their Gini importance rankings.

*Transmission Disequilibrium for MSNP1*

A SNP directly overlapping *MSNP1* (T allele at SNP rs4307059), the pseudogene for membrane-organizing extension spike protein, was identified as associated with ASD in the original study from which some of the data we used here were produced, but was not statistically significant (Ma et al., 2009). A second study in the same year with larger sample sizes and different approaches identified the same SNP as statistically significant (Wang et al., 2009). In that study, the authors performed a pedigree disequilibrium test on 780 families from the Autism Genetic Resource Exchange cohort genotyped at 486,864 SNPs. No significant loci were found at the standard significant *p*-value (*p* < 5 x 10^-8^). They then performed a standard GWAS with 1,204 cases and 6,491 controls from CHOP (Children’s Hospital of Philadelphia) and also found no significant loci. Next, they combined datasets from multiple sources and imputed SNPs, which identified a significant association with the rs4307059 SNP (*p* < 3.4 x10^-8^). Replication of this SNP association was attempted with a third data set from 1,390 cases from 447 families genotyped at 1 million SNPs (some of the data used for this study). The associations were in the same direction but not significant. A fourth data set of 108 cases and 540 controls genotyped at 300,000 SNPs confirmed the rs4307059 locus (*p* < 2.3 x 10^-10^) but with imputed SNPs. They also attempted to identify CNV with the PennCNV (Wang et al., 2007) approach and found several CNVs but because they were already listed in the Database of Structural Variants, they assumed they were not causative of ASD.

It is important to note that the quality control for Wang et al. 2009 included the removal of SNPs in which NMI was detected in more than 5% of the ASD individuals. They also excluded families in which more than 2% of loci demonstrated NMI. In addition, they removed SNPs that did not conform to HWE (p < 0.001) and assume that “*Samples with excessive Mendelian errors could indicate potential paternity problems, sample mislabeling, or sample handling problem during the genotyping experiments, and should be excluded from downstream association analysis*”. Under these criteria, the majority of the ASD-SV_NMI_ we identified in this study were removed and most likely rs11739167 would have been removed because it violated both HWE and MI.

The *MSNP1* locus produces an antisense transcript that is 94% identical to the moesin (*MSN*) mRNA located on the X chromosome and it has been shown to reduce MSN protein levels when overexpressed *in vitro* (Kerin et al., 2012). That same study found 12.7-fold more mRNA for *MSNP1* and 2.4-fold more for *MSN* in ASD post-mortem brain tissue (temporal cortex) compared to controls. Genotypes from rs7704909 (T/T), rs12518194 (A/A), and rs430705 (T/T) were positively correlated with mRNA expression of *MSNP1*. The two cadherin genes flanking *MSNP1* were not different amongst these genotypes, suggesting that they are not associated with the molecular aspects of ASD at this locus. Paradoxically, the MSN protein was not different in the temporal cerebral cortex of ASD post-mortem tissue compared to controls even though mRNA levels differed significantly. The authors suggested that the knockdown of MSN is important only during early development, which is supported by work that showed that expression peaks near birth and declines postnatally. Overall, independent expression data supports the involvement of *MSNP1* in ASD.

MSN is a component of a multi-protein complex of proteins called ERM (ezrin/radixin/moesin) that links cytoskeletal proteins to the membrane and have been heavily implicated in neurological processes. MSN is expressed at the leading edge of the growth cone of migrating neurons and it is phosphorylated at a threonine (T558) via AMPA, NMDA, and mGluR5 glutamate receptor signaling (see section below on glutamate receptors) (Kim et al., 2010). This appears to mechanistically occur via intracellular calcium release mediated by ryanodine receptors from the endoplasmic reticulum modulated by activation of mGluR5 (Breit et al., 2018). That same study found that glutamate induced phosphorylation of MSN is also necessary for synaptic vesicle trafficking. In addition, exposure of hippocampal neurons to glutamate induced activation of MSN and was associated with an increase in the number of active synaptic boutons, the presynaptic axon terminals that contact dendritic spines to form a synapse.

In order to determine cellular effects of *MSNP1* at the molecular level, the group that originally reported differential expression in post-mortem brain tissue overexpressed *MSNP1* mRNA in neural progenitor cells (DeWitt et al., 2016a), which caused decreased neurite outgrowth (number and length) *in vitro* and caused changes in gene expression although none survived Bonferroni correction. Although not statistically significant, a Gene Ontology analysis of the differentially expressed genes (DEGs) that were significant from uncorrected *p*-values (*p* < 0.05) found enrichment for chromatin remodeling and regulation, including histones and the SWI/SNF pathway. The same lab carried out a knockout of *MSNP1* in the same cell lines used in the over-expression study and although they found no change in *MSN* mRNA, they did identify roughly 1,300 DEGs that also included chromatin remodeling and the immune response (DeWitt et al., 2016b). The most differentially regulated gene was OAS2 (increased 150-fold), which is an immune-related gene that is associated with ADHD (Jong et al., 2016). All of these results mechanistically link *MSNP1* to SVs of large effect that we detail below.

The paradox of no correlation between *MSNP1* and protein levels of MSN in post-mortem brain tissue of ASD individuals was addressed by a study that used an enhancer trap approach to determine the functionality of other sequences in the 100kb region containing *MSNP1* and linked to rs4307059. They generated transgenic mice expressing a beta-gal reporter from a BAC clone containing the rs4307059 risk allele along with controls that contained portions of the 100kb region (Inoue and Inoue, 2016). Only the BAC with the risk alleles and *MSNP1* demonstrated LacZ expression in the mouse cerebral cortex, striatum, and cerebellum; the other BACs with the non-risk alleles did not. Importantly, this pattern was not present when the paralogous region from the mouse was used, indicating that this region is human- or primate-specific. More specifically, the enhancer activities were localized in the S1 cortical layer II/III, striatal, and cerebellar neurons and consistent with the post-mortem results above, they showed no change in the *MSN* mRNA or protein.

Further support of the role of *MSNP1* in ASD was found with a study on 7,313 children (86 of whom were diagnosed with ASD). The rs4307059 SNP was associated with decreased social communication phenotypes and the strongest single-trait associations were observed for stereotyped conversation and pragmatic communication skills (St Pourcain et al., 2010). Finally, dysregulation of MSN in early development appears to alter long term memory in a *Drosophila* model that was independent of its role in development (Freymuth and Fitzsimons, 2017).

In order to interrogate the rs11739167 SNP in greater detail, we re-plotted the SNP genotypes (array intensity values) for the fathers, mothers, and ASD offspring separately (Fig. 2 in the main text and Fig. S3). Unexpectedly, the genotypes called as “-/-” in the fathers appear to be a sub-group of heterozygotes. The expectation was that there would be no signal at all for “-/-” as seen with NRXN3. The mothers did not display this subtype, but notably the plot revealed a shift *upwards* in the signal intensity for the G allele in all females. The most likely explanation is that the Illumina probe for rs11739167 is binding to the paralogous sequence in exon 3 of the *MSN* gene on the X chromosome (the sequences only differ by a single base), which produces two extra “G” calls in females and an extra “G” call in males because of their hemizygosity. This explains the increased signal in females for the G allele. However, ***this does not explain the shift of the subset of males on the x-axis*** because a probe that binds to the X chromosome *MSN* gene will ***only*** produce a “G”, which is defined by the y-axis, i.e., there is no SNP in the *MSN* gene at this location, there is only a variable SNP site in the *MSNP1* gene on chromosome 5. Therefore, ***a shift in the X-axis is autosomal and not sex linked***. We plotted the remaining 13 transmission disequilibrium tests (TDT) SV_NMI_ in the same manner as *MSNP1* to determine if they are identifying true SV and/or pseudogenes. The majority identify CNV and two appear to be artifacts from binding to a pseudogene copy (Fig. S3).

**Supplementary Text**

*Neurexin-3 NMI*

The SNP rs221465 in the NRXN3 gene displays NMI in 35% of ASD individuals. This site is proximal to a ncRNA near an intron/exon border, a histone methylation site, and an enhancer that is expressed during neural tube development, making it an attractive candidate for ASD association. However, the most recent version of the human genome reported an 8.6 kb deletion at this location with an allele frequency of 0.28. After we re-scored the genotypes for this deletion in the GWAS population using the combination of raw intensity values and parental inheritance, we found normal Mendelian inheritance, conformation to Hardy-Weinberg Expectations, and no statistical difference from the 1000 Genome EUR population. This suggests that this SV is a false positive in the context of ASD, but also confirms that NMI is an accurate means to identify SVs based on information of normally segregating variants in the 1000 Genome population.

**Glutamate Signaling**

Our Gene Ontology analysis of the SV in coding regions identified several categories associated with glutamate signaling (Table S6a). Disrupted glutamate signaling has been thoroughly described in ASD [[34]](https://paperpile.com/c/0BBqdN/Fp688) and in the ASD-like Kleefstra Syndrome [[72]](https://paperpile.com/c/0BBqdN/pCodn). Glutamate receptors mediate excitatory synapse transmission in the brain and were originally classified according to the glutamate analogs they bound (*23*, *24*). There are five families of receptors, all of which have been implicated in ASD. Four of the five function as transmembrane ion channels; these are known as ionotropic glutamate receptors or iGluRs. The fifth type are the metabotropic G-protein coupled glutamate receptors (mGluRs) and unlike the iGluRs, they respond through classic signal transduction pathways. All of these receptors are an important component of cerebellum function and development.

Even though the cerebellum comprises only 1/10^th^ of the total brain volume, it is the most dense region and contains more neurons than the rest of the brain combined [[38,73]](https://paperpile.com/c/0BBqdN/LdqS5+iaUpw). Although this brain structure is most commonly associated with motor skills and physical movement, it also functions in the accurate *coordination* of motor skills as well as language processing and expression of emotion [[39,40]](https://paperpile.com/c/0BBqdN/ALfRA+XpGPY). Damage to different regions of the cerebellum results in impaired communication similar to ASD and cerebellar injury at birth increases the diagnosis of ASD by 36-fold [[40]](https://paperpile.com/c/0BBqdN/XpGPY). The cerebellum rapidly grows during the third trimester of pregnancy and differentiates early in development, but it is not mature until the first postnatal years [[39]](https://paperpile.com/c/0BBqdN/ALfRA). A highly organized network resides in the cerebellum that is composed of Climbing Fibers, each of which is connected to a single Purkinje Fiber that integrates into an orthogonal layer of Parallel Fibers (composed of granule cells) through many synapses. Nearly all post-mortem examinations of ASD brains have identified differences in the cerebellum compared to controls [[74]](https://paperpile.com/c/0BBqdN/sZb1) , and the most consistent observations are the loss of Purkinje Fiber cells, overall cerebellar enlargement early in development, and reduction in size by adulthood [[39,73]](https://paperpile.com/c/0BBqdN/ALfRA+iaUpw). Functional differences of the cerebellum among ASD individuals are also widely reported. Although we identify SV in all types of glutamate receptors and accessory proteins, the frequency of SV and the subunits affected strongly implicate the cerebellum in ASD. We summarize each of the categories below.

*AMPAR - α-amino-3-hydroxy-5-methyl-4-isoxazolepropionic acid receptor*

The majority of fast excitatory synaptic transmission in the mammalian central nervous systems is mediated by AMPA receptors that are heterodimers of one of the four subunit types (GRIA1-4)[[32]](https://paperpile.com/c/0BBqdN/DglW). These receptors are also important for NMDA-modulated plasticity and as with other glutamate receptors, splice variants and different combinations of heterodimers produce a diversity of receptor types [[75]](https://paperpile.com/c/0BBqdN/3CeA6). AMPA typically modifies NMDA signaling by releasing voltage-dependent activity-blocks from extracellular Mg^2+^ to those receptor types [[76]](https://paperpile.com/c/0BBqdN/6enKY). The GRIA2 subunit is unusual in that it undergoes RNA-editing, which directly affects the permeability of the channel pore itself and is the major form found in the adult brain [[77]](https://paperpile.com/c/0BBqdN/N3y7T). The majority of heterodimers of these receptors are composed of GRIA1 and 2 but GRIA4 is expressed highly in the developing neonatal brain[[78]](https://paperpile.com/c/0BBqdN/P4Fhf) and in the adult it is mainly found in the cerebellum as a homodimer in Bergmann’s Glia [[32,78]](https://paperpile.com/c/0BBqdN/P4Fhf+DglW) (see GluD below) or interneurons[[77]](https://paperpile.com/c/0BBqdN/N3y7T). Deletion of the GRIA4 subtypes in these cells in young mice results in the disruptions between granule cells of the Parallel fiber layer and Purkinje cells.

Overall, we find that ASD cases have SVs in several *GRIA* subunits (Table S8). As with all glutamate receptors, AMPAR have numerous accessory subunits that participate in presentation and signaling that include the stargazing family of proteins (CACNG1-8), the SHISA family of proteins, as well as IL1RAP1L, GRIP1 and GRIP2, and the tyrosine phosphatase PTPRD that binds to IL1RAPL1 [[79]](https://paperpile.com/c/0BBqdN/vppVR). Several of these have been associated with ASD in other work and display ASD-SV (Table S8). Just under 15% of cases display an SV in CACNG2, which results in loss of excitatory transmission between mossy fibers and granule cells of the Parallel Fibers when deleted [[75]](https://paperpile.com/c/0BBqdN/3CeA6) .

*NMDAR - N-methyl-D-aspartate receptor*

At most synapses, NMDA and AMPA are expressed at postsynaptic membranes and are co-activated by glutamate secreted from the presynaptic terminal. As with the other glutamate receptors, NMDA exists as multimers of different subunits, although all contain at least one GRIN1 subunit and usually GRIN2[[32]](https://paperpile.com/c/0BBqdN/DglW). In our analysis, many ASD cases carry an ASD-SV in at least one NMDA subunit as well as several supporting proteins for NMDA function. The majority of individuals harbor an SV in the KALRN gene, which is necessary for NMDA-dependent plasticity [[80]](https://paperpile.com/c/0BBqdN/Elja5). We did not detect an ASD-SV in the obligatory *GRIN1* subunit, which may indicate strong purifying selection for proper function. The two subunits demonstrating the highest levels of ASD-SV (*GRIN3A* and *GRIN2B*), as with other SV-containing glutamate receptor subunits discussed here, are important for early postnatal development [[32]](https://paperpile.com/c/0BBqdN/DglW). Nearly 1/3 of individuals carry ASD-SV in *GRIN3A*, which alters NMDA signaling in a dominant negative manner when present[[81]](https://paperpile.com/c/0BBqdN/O6TP5). As GRIA4, GRIN3A is specific to and important for early brain development, which includes expression in astrocytes (e.g., Bergmann’s glia). Finally, physical activity regulates expression of GRIN2B in cerebellum granule cells (Parallel Fibers)[[32]](https://paperpile.com/c/0BBqdN/DglW).

*KAR – Kainate receptor*

KAR are unlike the other glutamate receptors in that they tend to modulate or regulate the synaptic activity of the other types and regulate neurotransmitter release [[82]](https://paperpile.com/c/0BBqdN/5fOuj). They are also necessary for a unique NMDA-independent form of plasticity in the hippocampus [[83]](https://paperpile.com/c/0BBqdN/uFWeS), an area that shows decreased activity in ASD and is linked to short term memory [[84]](https://paperpile.com/c/0BBqdN/RCky0). Loss of function mutations in the GRIK2 subunit cause severe intellectual disability and appear to be responsible for mood disorders. KARs differ from NMDAR and AMPAR in that they can be present at both pre- and postsynaptic membranes[[32]](https://paperpile.com/c/0BBqdN/DglW). KAR have been shown to modulate synaptic transmission at mossy fiber-CA3 pyramidal cells, which feed directly to Purkinje cells in the cerebellum (GluD below). Many ASD cases carry an ASD-SV in at least one *GRIK* subunit of KARs with the majority occurring in *GRIK2*, a gene that has been associated with ASD in several other studies (Table S12).

The most frequent ASD-SV site overlaps and is identified by the SNP rs2051449. This site resides 600 base pairs from a ChIP-Seq site for PCBP2, SRSF9, and HNRNPK, all of which participate in RNA-splicing. It is therefore likely that this ASD-SV disrupts proper splicing of the adjacent exon 12 of the gene. This likely results in the loss of exon 12, directly affecting the glutamate binding pocket. It is possible that the exon-depleted form of KAR assembles but does not signal, producing a dominant negative phenotype.

*GluD – Glutamate Receptor Delta*

GluD receptors are an important component of the neurobiology of the cerebellum. There are two GluDs (GLUD1 and GLUD2 proteins encoded by *GRID1* and *GRID2* genes, respectively). GluD2 binds serine as well as a family of proteins called cerebellins (Cblns), which are secreted from granule cells onto Purkinje Fiber cells with the assistance of the Bergmann’s Glia [[85]](https://paperpile.com/c/0BBqdN/Z6WLO). The highly organized network of the cerebellum is disrupted in GRID2 knockout mice in several ways; rather than a single Climbing Fiber cell connecting to a single Purkinje Fiber cell, Climbing Cells connect to numerous Purkinje Cells and granule cells that comprise the Parallel Fibers in the orthogonal layer. It appears that these connections are meant to be pruned during brain development and the loss of GRID2 prevents this. In addition, AMPA receptors are expressed at much higher levels in GRID2 knockout mice than wildtype mice, suggesting that a normal function of GRID2 is to suppress AMPA expression [[86]](https://paperpile.com/c/0BBqdN/GN9di). Unlike the other four glutamate receptors, GluDs do not directly bind glutamate. Most ASD individuals carry an ASD-SV in the GRID2 gene.

*mGLURs – Metabotropic glutamate receptors*

Unlike the other glutamate receptors, metabotropic glutamate receptors (mGLURs) are G-protein coupled receptors (GPCRs) that signal through a traditional intracellular cascade upon binding ligand instead of acting as an ionic channel as the other receptors do. mGLURs also exist as dimers rather than tetramers as most iGLURs. The eight known mGLURs are divided into three groups based on intracellular signaling and biological effect. Group 1 (GRM1 and GRM5) act to release intracellular calcium stores for propagation of signal whereas those in Groups 2 (GRM2 and GRM3) and Group 3 (GRMs 4,7, and 8) act through adenylate cyclase. These latter two groups also inhibit the release of the inhibitory neurotransmitter GABA (gamma-aminobutyric acid)[[87]](https://paperpile.com/c/0BBqdN/jY7Yc). More than half of cases harbor an SV in one of the Group I mGluRs, with the highest in GRM5. As with many of the other ASD-associated glutamate receptor subunits in this study, GRM5 is expressed early in development in Purkinje Fibers and declines into adulthood [[88]](https://paperpile.com/c/0BBqdN/jRgqg). GRM5 has been shown to immunoprecipitate and function with GluD1 (see GluD above), which results in altered AMPA expression[[89]](https://paperpile.com/c/0BBqdN/LH83b). GRM1 and GRM5 also interact with NMDA receptors via DLG4, SHANK, and HOMER proteins[[90]](https://paperpile.com/c/0BBqdN/BmPWP), which have been implicated in ASD and function as associated proteins with GluDs. Finally, GRM5 has been shown to be a necessary component of AMPA/NMDA-mediated phosphorylation of moesin for dendritic spine development and axon guidance [[36]](https://paperpile.com/c/0BBqdN/Qn4xA).

**Axon Guidance**

The development of complex neural circuits requires the migration of axons over long distances to make the appropriate connections to their target cells. This process requires an axon guidance “cone” at the tip, which senses attractant or repulsive cues secreted by astrocytes and other cells that lie along the path. The axons turn based on the combination of the molecule secreted and the receptor(s) being expressed at the tip of the cone. Upon passing a secreting sentinel cell, the receptors at the tip are degraded and replaced with new receptors that will sense the next decision point in the pathway. Often the axon will make contacts with the cell it passes via contactin and contactin-associated proteins (CNTNs and CNTNAPs) that, as mentioned above, are part of the NCAM-associated SVs.

The majority of the axon-guidance related genes harboring ASD-SV are either the receptors expressed at the cone of the migrating axon or their partner ligand that is secreted by the cells at the choice point. The two most affected pairs are the *Netrin/DCC* and the *ROBO1/SLIT1* genes followed by NRP1 and the Semaphorins (Table S17). The largest group of axon guidance genes affected are the Ephrin receptors, which are heavily involved in the development of the superior colliculus[[50]](https://paperpile.com/c/0BBqdN/lQvsE), notably knockout mice of EPHA8 fail to develop proper connections within this structure (OMIM #176945). The superior colliculus functions to initiate behavioral responses to visual cues in the external world [[91]](https://paperpile.com/c/0BBqdN/eJYkp).

### **Supplementary Figures**

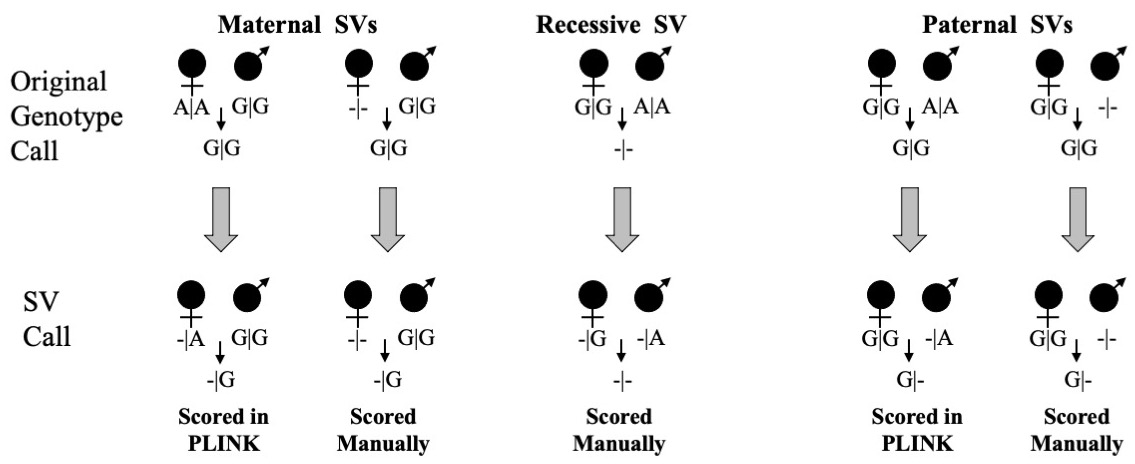

**Figure S1.** Overview of SV detection using NMI. Genotypes are assembled from family pedigrees and scored using standard Mendelian tests, such as those used in the software PLINK, and will identify maternal vs paternal NMI. PLINK results are represented by A|A G|G parents and G|G offspring for maternal NMI (top left family) and G|G A|A parents with G|G offspring (second family from the far right). We added scenarios where no genotype is called (-/-), which is ignored by PLINK but that likely represent a full deletion, whereas homozygous offspring from NMI families likely represents heterozygous deletion or haploinsufficiency (e.g., -|G). In order to identify ASD-specific SVs, we filter out any NMI that overlaps a known SV from the 1000 Genome project and other available data sets. We also overlapped our potential ASD-SVs with the latest dbSNP build to remove any that overlap a SNP or INDEL with a minor allele frequency greater than 0.02 because these may cause aberrant probe binding and essentially act like an SV, but the pattern is not specific to ASD, rather it is simply identifying common non-ASD variation due to underlying cryptic SNPs that disrupt the genotyping assay.

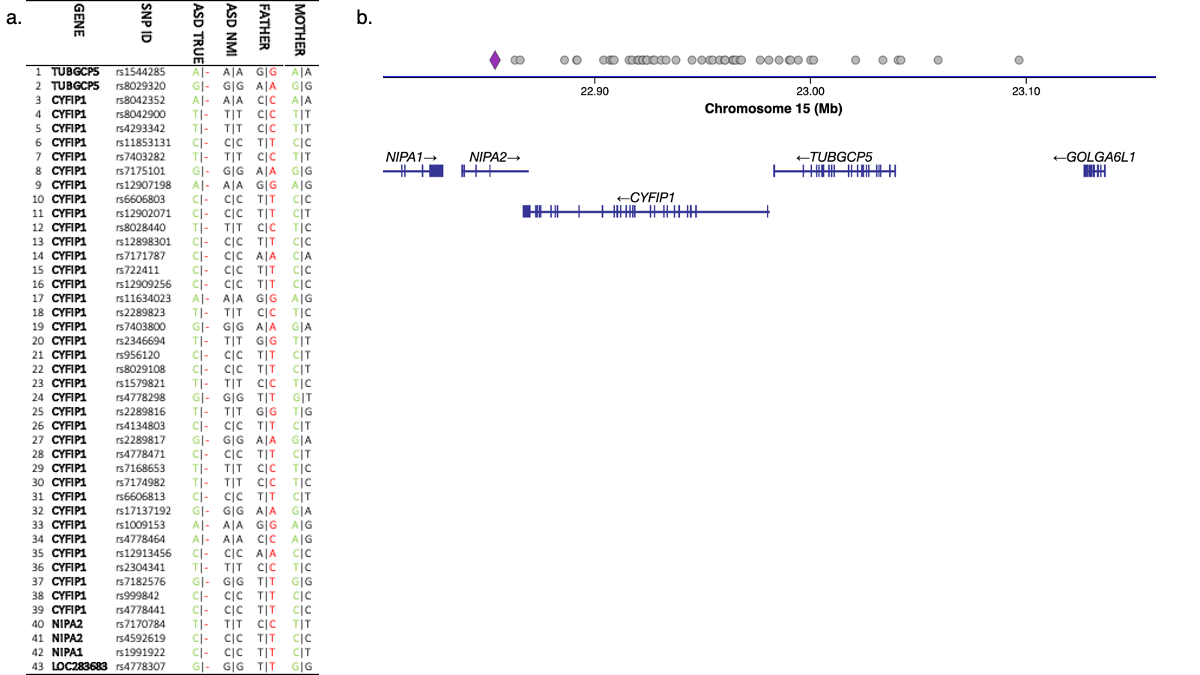

**Figure S2.** Examples of large SV detected by a long run of 43 NMI. (a) The probability of finding 43 sequential NMI SNPs based on the overall error rate given the binomial distribution described above is 1.2 x 10^-105^(i.e., the probability of obtaining 43 successes in 43 trials given an overall error rate of 0.36% where a success is defined as an NMI pattern). Likely, the paternal alleles are deleted (red nucleotides) and the child is hemizygous for the maternal haplotype (green nucleotides), consistent with published models. (b) This particular run of NMI SNPs identifies a well characterized region on chromosome 15 that causes neurodevelopmental disorders including Angleman’s Syndrome. Our method therefore represents a means to rapidly identify these individuals (even if they are rare) as we have done here with two different datasets.

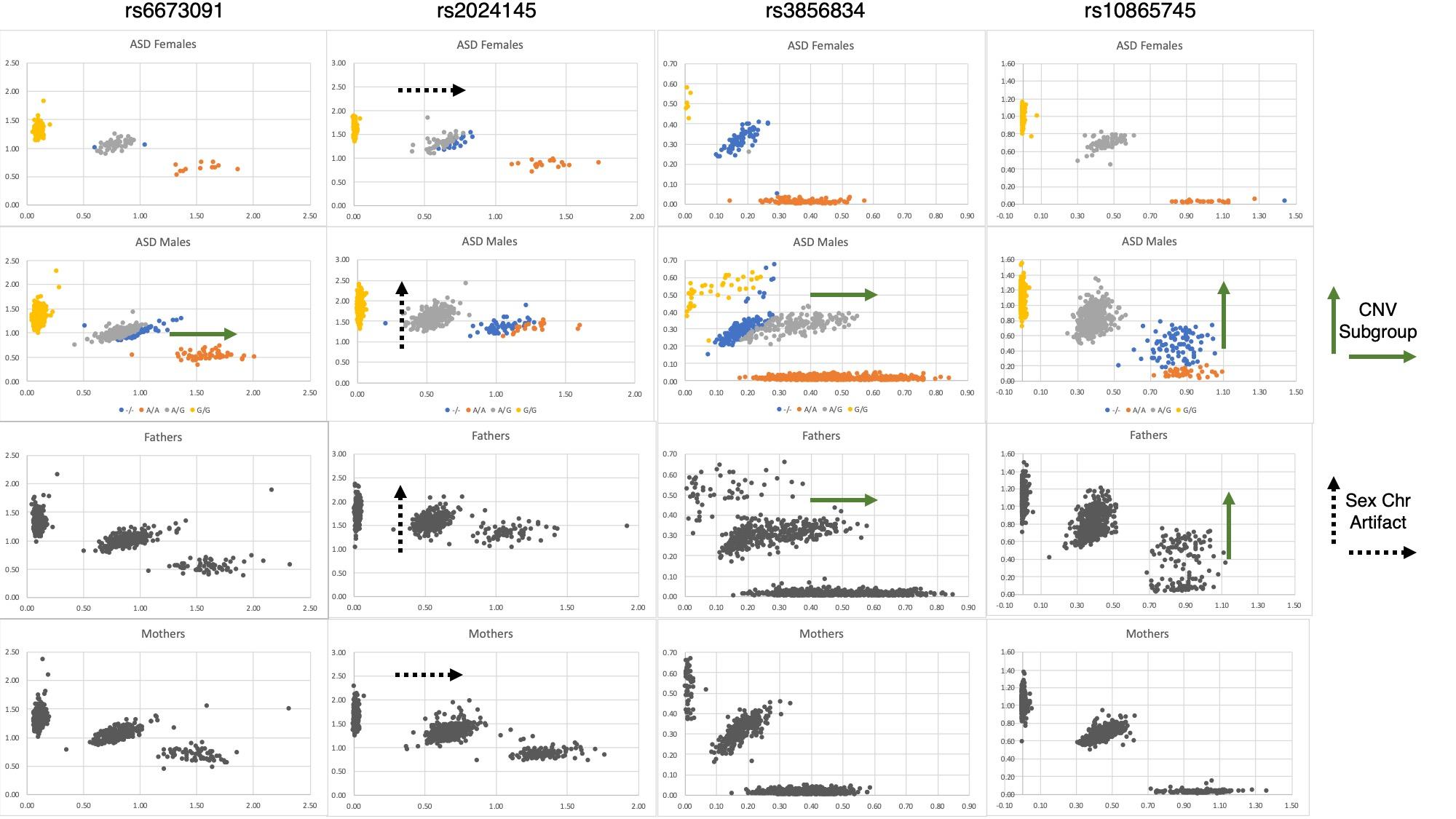

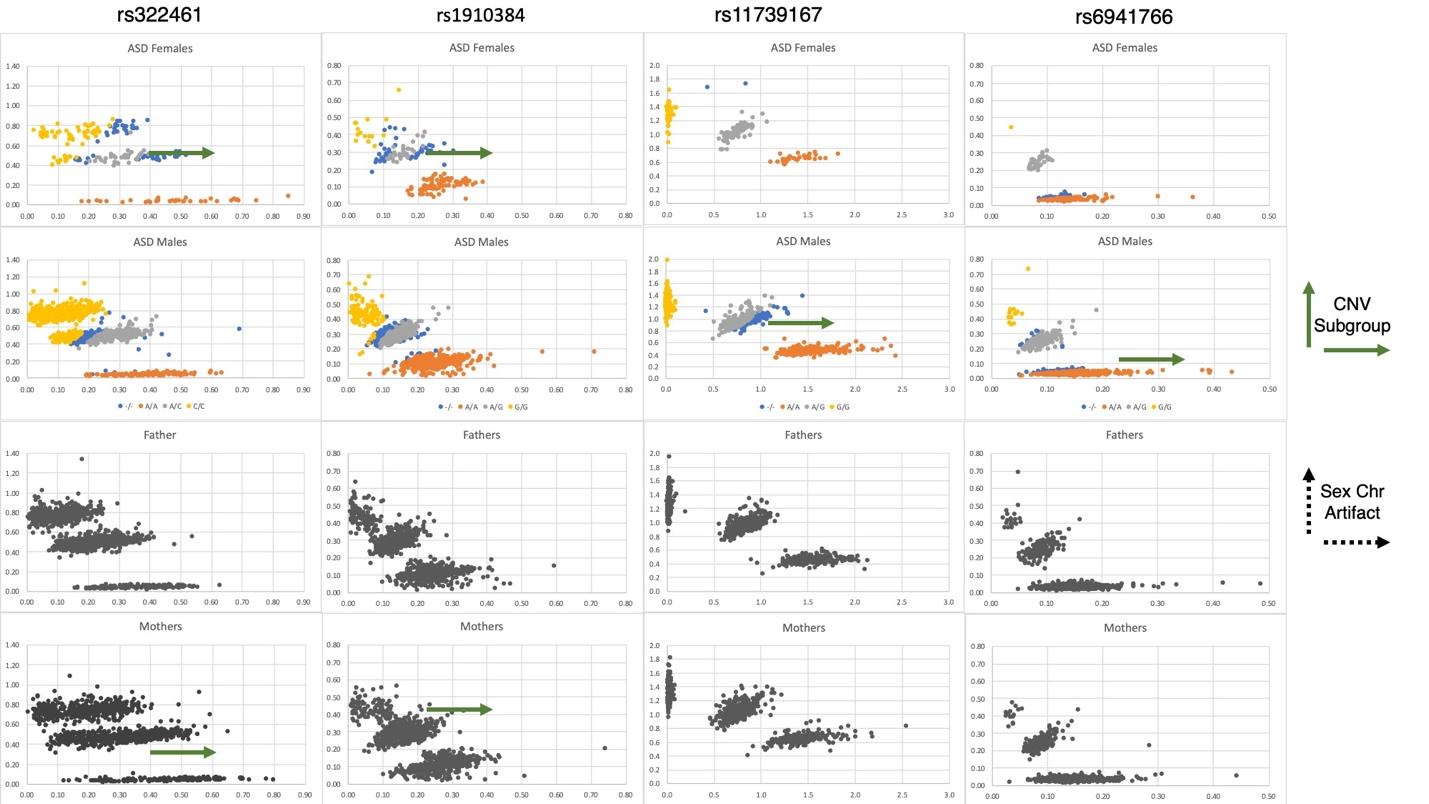

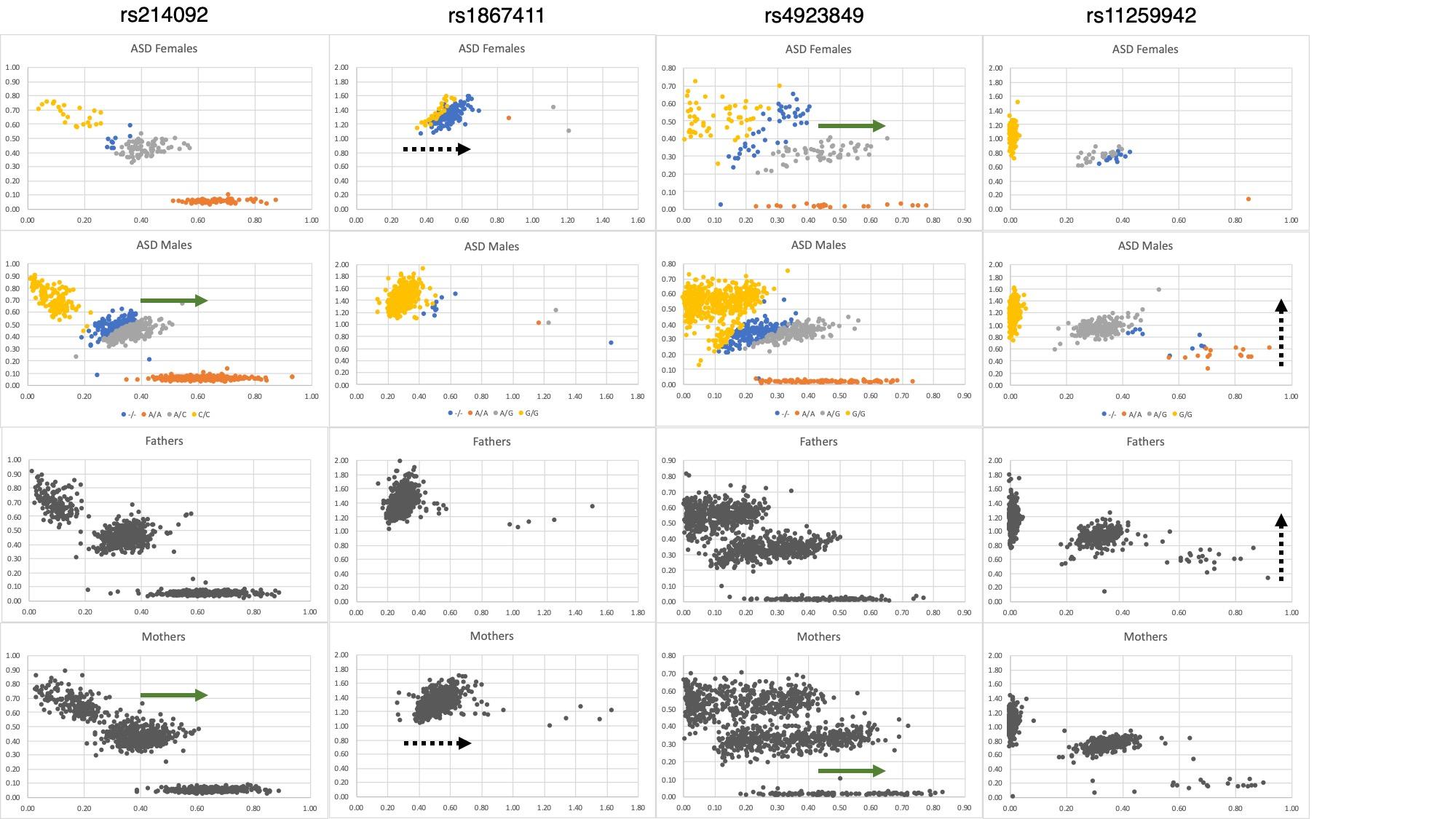

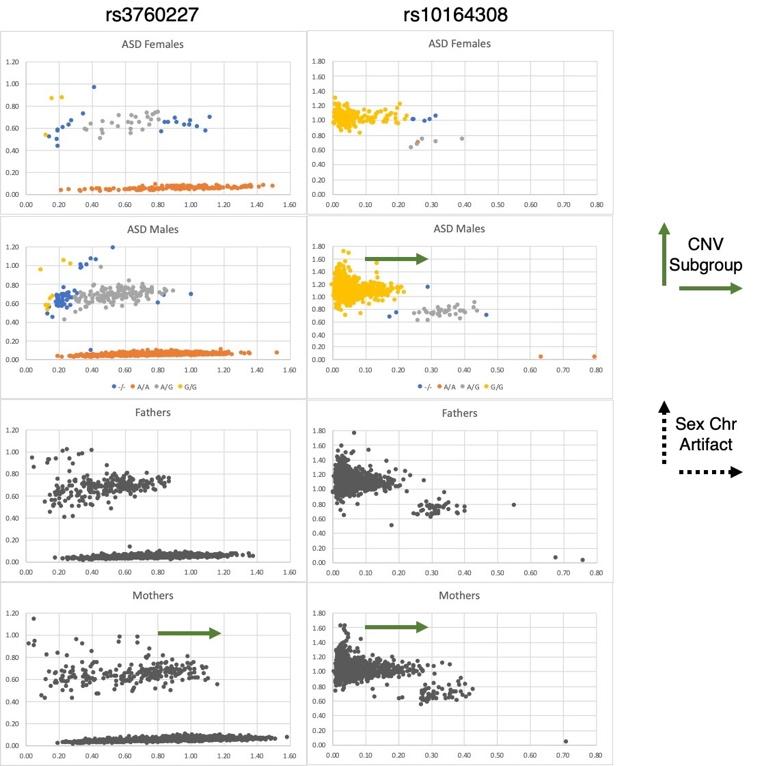

**Figure S3. Detailed plots of ten maternal and four paternal TDT for fathers, mothers, and offspring.** Eleven of the fourteen TDTs identify SV/CNV in a sub-group of individuals and three are likely binding to pseudogenes on sex chromosomes because either all males (Y chromosome pseudogene) or all females (X chromosome pseudogene) shift on the axis (rs2024145, rs1867411, rs11259942, broken arrows). Two of the loci identify the same known CNV (nsv4046446) on the Y chromosome (rs3856834 and rs10865745). Their frequency does not differ from non-ASD individuals and is therefore not associated with ASD. One of them, rs10164308, resides in a *HERK11* transposon, which is known to be active in the human genome and therefore this CNV gain may represent the movement of that fragment to different positions in the genome. Further study is needed to determine if this and the other loci are associated with ASD. Several lines of evidence implicate *MSNP1,* namely: (1) it is centered on the original GWAS signal from the study that produce the data, (2) *in vitro* experiments determined that it regulates the expression and translation of the parent gene *MSN*, (3) it is differentially expressed in post-mortem brain tissue from ASD and non-ASD individuals, (4) it participates directly in dendritic spine formation as a large multi-subunit complex that includes other known ASD-linked genes such as *CYFIP1* in the WAVE complex, and (6) it is linked to two other ASD-SV_NMI_ (*GRIK2* and *PTPRD*) in a GWAS for Obsessive Compulsive Disorder.
